# Trends, variation and clinical characteristics of recipients of antivirals and neutralising monoclonal antibodies for non-hospitalised COVID-19: a descriptive cohort study of 23.4 million people in OpenSAFELY

**DOI:** 10.1101/2022.03.07.22272026

**Authors:** The OpenSAFELY Collaborative, Amelia Green, Helen J Curtis, Rose Higgins, Rebecca Smith, Amir Mehrkar, Peter Inglesby, Viyaasan Mahalingasivam, Henry Drysdale, Nicholas J DeVito, Richard Croker, Christopher T Rentsch, Krishnan Bhaskaran, Colm Andrews, Seb Bacon, Simon Davy, Iain Dillingham, David Evans, Louis Fisher, George Hickman, Lisa Hopcroft, William J Hulme, Linda Nab, Jon Massey, Orla McDonald, Jessica Morley, Caroline E Morton, Robin Park, Alex J Walker, Tom Ward, Milan Wiedemann, Chris Bates, Jonathan Cockburn, John Parry, Frank Hester, Sam Harper, Ian J Douglas, Stephen JW Evans, Laurie Tomlinson, Brian MacKenna, Ben Goldacre

## Abstract

**Objectives:** Ascertain patient eligibility status and describe coverage of antivirals and neutralising monoclonal antibodies (nMAB) as treatment for COVID-19 in community settings in England.

**Design:** Cohort study, approved by NHS England.

**Setting:** Routine clinical data from 23.4m people linked to data on COVID-19 infection and treatment, within the OpenSAFELY-TPP database.

**Participants:** Non-hospitalised COVID-19 patients at high-risk of severe outcomes.

**Interventions:** Nirmatrelvir/ritonavir (Paxlovid), sotrovimab, molnupiravir, casirivimab or remdesivir, administered in the community by COVID-19 Medicine Delivery Units.

**Results:** We identified 102,170 non-hospitalised patients with COVID-19 between 11^th^ December 2021 and 28^th^ April 2022 at high-risk of severe outcomes and therefore potentially eligible for antiviral and/or nMAB treatment. Of these patients, 18,210 (18%) received treatment; sotrovimab, 9,340 (51%); molnupiravir, 4,500 (25%); Paxlovid, 4,290 (24%); casirivimab, 50 (<1%); and remdesivir, 20 (<1%). The proportion of patients treated increased from 8% (180/2,380) in the first week of treatment availability to 22% (420/1870) in the latest week. The proportion treated varied by high risk group, lowest in those with Liver disease (12%; 95% CI 11 to 13); by treatment type, with sotrovimab favoured over molnupiravir/Paxlovid in all but three high risk groups: Down syndrome (36%; 95% CI 31 to 40), Rare neurological conditions (46%; 95% CI 44 to 48), and Primary immune deficiencies (49%; 95% CI 48 to 51); by ethnicity, from Black (10%; 95% CI 9 to 11) to White (18%; 95% CI 18 to 19); by NHS Region, from 11% (95% CI 10 to 12) in Yorkshire and the Humber to 23% (95% CI 22 to 24) in the East of England); and by deprivation level, from 12% (95% CI 12 to 13) in the most deprived areas to 21% (95% CI 21 to 22) in the least deprived areas. There was also lower coverage among unvaccinated patients (5%; 95% CI 4 to 7), those with dementia (5%; 95% CI 4 to 6) and care home residents (6%; 95% CI 5 to 6).

**Conclusions:** Using the OpenSAFELY platform we were able to identify patients who were potentially eligible to receive treatment and assess the coverage of these new treatments amongst these patients. Targeted activity may be needed to address apparent lower treatment coverage observed among certain groups, in particular (at present): different NHS regions, socioeconomically deprived areas, and care homes.

**What is already known about this topic:** Since the emergence of COVID-19, a number of approaches to treatment have been tried and evaluated. These have mainly consisted of treatments such as dexamethasone, which were used in UK hospitals,from early on in the pandemic to prevent progression to severe disease. Until recently (December 2021), no treatments have been widely used in community settings across England.

**What this study adds:** Following the rollout of antiviral medicines and neutralising monoclonal antibodies (nMABs) as treatment for patients with COVID-19, we were able to identify patients who were potentially eligible to receive antivirals or nMABs and assess the coverage of these new treatments amongst these patients, in as close to real-time as the available data flows would support. While the proportion of the potentially eligible patients receiving treatment increased over time, rising from 8% (180/2,380) in the first week of the roll out to 22% (420/1870) in the last week of April 2022, there were variations in coverage between key clinical, geographic, and demographic subgroup.

**How this study might affect research, practice, or policy:** Targeted activity may therefore be needed to address lower treatment rates observed among certain geographic areas and key groups including ethnic minorities, people living in areas of higher deprivation, and in care homes.

## BACKGROUND

Since the emergence of COVID-19, a number of approaches to treatment have been tried and evaluated^1^. In UK hospitals, treatments such as dexamethasone were used from early in the pandemic to prevent progression to severe disease^2^. However, no treatments were widely used in the community, where care was largely supportive and focussed on detection of need for hospital admission^3^. In April 2021, the UK government established a Therapeutics and Antivirals Taskforce with the aim of identifying and deploying new medicines to treat COVID-19 in community settings to reduce the risk of hospital admission^4^.

On 16^th^ December 2021, new COVID Medicine Delivery Units (CMDUs) were launched across England, offering antiviral medicines and neutralising monoclonal antibodies (nMABs) as treatment to patients with COVID-19 at high risk of severe outcomes in outpatient clinics or their own home^5^. Initially sotrovimab, casirivimab/imdevimab, and molnupiravir were available at these units with nirmatrelvir/ritonavir (Paxlovid) and remdesivir becoming available in February 2022. The UK government established an expert clinical group to develop criteria to support identification of high risk groups eligible for these treatments using NHS data^6^. The NHS in England issued detailed clinical commissioning policies^7^ and people identified as high risk were informed by letter that in the event that they test positive for SARS-CoV-2 they would be eligible for these treatments.

OpenSAFELY is a secure analytics platform for electronic patient records built by our group on behalf of NHS England to deliver urgent academic and operational research during the pandemic^8,9^. Analyses can currently run across all patients’ full pseudonymised primary care records, with patient-level linkage to various sources of secondary care data. Data on patients receiving antivirals and nMABs from CMDUs was similarly linked and is now updated weekly with approximately one week’s lag time. Code and analysis is shared openly for inspection and re-use.

This study set out to identify patients registered with OpenSAFELY-TPP practices who were potentially eligible to receive antivirals or nMABs in a community setting and assess the coverage of these new treatments amongst these patients, in as close to real-time as the available data flows would support. We also describe how coverage varied between key clinical, regional, and demographic subgroups, and whether any treatments given may have been potentially inconsistent with guidance.

## MATERIALS AND METHODS

### Study design

We conducted a retrospective cohort study beginning on the 11^th^ December 2021 (the earliest date that a patient could have tested positive to be eligible for receiving treatment when they became available from CMDUs from 16^th^ December 2021) and ending on 28^th^ April 2022. Regular treatment coverage reports have also been produced and updated regularly with extended follow-up time using near real-time data as the treatment programme progresses^10^.

### Data sources

This analysis was conducted using the OpenSAFELY-TPP platform which executes code across records for all patients currently registered with general practices using TPP SystmOne electronic health records (EHR) software: this is approximately 23.4 million people, or 40% of the English population. It includes pseudonymised data such as coded diagnoses, medications and physiological parameters. No free text data are included. This primary care data is linked, via hashed NHS numbers, to: accident and emergency (A&E) attendance and in-patient hospital spell records via NHS Digital’s Hospital Episode Statistics (HES); national coronavirus testing records via the Second Generation Surveillance System (SGSS); and the “COVID-19 therapeutics dataset”, a patient-level dataset on antiviral and nMAB treatments, newly sourced from NHS England, derived from Blueteq software that CMDUs use to notify NHS England of COVID-19 treatments. Vaccination status is available in the GP records directly via the National Immunisation Management System (NIMS).

### Study population

We included all individuals aged 12 or over with a positive SARS-CoV-2 test on or after 11^th^ December 2021 and all patients with a treatment record on or after 16^th^ December 2021, who were registered with a GP at the time of their test/treatment.

### Eligibility for treatment

We identified the population who were potentially eligible for treatment as those meeting the eligibility criteria for COVID-19 antiviral or nMAB treatment in the community: being a member of a high risk group (described below under High risk groups) and with confirmed SARS-CoV-2 infection (Box 1). There were two main differences to the official criteria in our implementation. Firstly, prior to 10th February 2022, infection should have been confirmed by a polymerase chain reaction (PCR) test (this was then relaxed to include lateral flow tests). We were not able to distinguish between lateral flow and PCR tests in all test records, and therefore included all positive SARS-CoV-2 test results across the whole study period. Secondly, having symptomatic COVID-19 was part of the eligibility criteria: however due to difficulties in determining symptom status (i.e. it was only possible to determine whether a patient’s positive test had a “symptomatic” flag at the time of the test, but not whether symptoms developed later) we did not implement this requirement in our analysis; we do however address this in a separate sensitivity analysis, where we restricted the potentially eligible population to only those with a “symptomatic” flag associated with their positive SARS-CoV-2 test to determine its use as an indicator of being potentially asymptomatic.

As we were unable to implement all of the eligibility criteria, patients who received treatment (see below) were also included in the population even if they were not identified as meeting eligibility criteria (e.g. having no positive SARS-CoV-2 test). All patients with records of more than one treatment in the community within two weeks of one another (potentially due to a data quality issue early in the rollout), or with an implausible treatment date (such as dates far into the future) were excluded due to not being able to accurately determine which treatment they received and when. The number of patients included/excluded for these reasons are reported.

**Box 1:** Eligibility and exclusion criteria, as per the Interim Clinical Commissioning Policy (published on 24^th^ February 2022)^7^ for non-hospitalised COVID-19 patients, and how these were applied in the present study. Earlier versions of the policy had some minor differences but this version was applied to the whole study period

| Eligibility/Exclusion Criteria |  | Criteria applied in present study* |
| --- | --- | --- |
| **Eligibility Criteria** | SARS-CoV-2 infection was confirmed by polymerase chain reaction (PCR) testing OR Lateral flow test (registered via gov.uk or NHS 119) | Positive PCR OR lateral flow SARS-CoV-2 test in SGSS. (As per NHS Digital's tests rule set logic<sup>6</sup>, patients with a prior positive SARS-CoV-2 within the last 30 days were excluded.) |
|  | Symptomatic with COVID-19 and showing no signs of clinical recovery | Not possible<sup>†1</sup> |
|  | Patient was a member of a 'high risk' group. | See Table 1 and Supplementary material Table S1 |
| **Exclusion criteria** | Requirement for hospitalisation for COVID-19 | Discharged from hospital within 30 days prior to positive test or treatment, where COVID-19 was the primary diagnosis. |
|  | New supplemental oxygen requirement specifically for the management of COVID-19 symptoms | Not possible |
|  | Known hypersensitivity reaction to the active substances or to any of the excipients of the medications below as listed in their respective Summary of Product Characteristics | Not possible |

\* Note: patients who received treatment (described below) were also included in the population even if they were not identified as meeting these criteria.
<sup>†1</sup> It was only possible to identify whether or not the patient's positive test had a "symptomatic" flag, but not whether symptoms developed later; therefore the symptomatic flag was used in a sensitivity analysis rather than within inclusion criteria.

### High risk groups

The detailed recommendations on the ten high risk groups derived by an expert clinical group (Box 2) were transformed by NHS Digital into detailed logic and analytic code, including a range of codelists, that could be used on datasets they held to identify all patients eligible for treatment by CMDUs^6^. However, NHS Digital recognised potential limitations in identifying all eligible patients using centrally held data, and freedom was permitted for CMDUs to use “non-digital” solutions to identify additional eligible patients locally based on the criteria outlined by the expert group. For example, patients with stage 4 chronic kidney disease would be eligible for treatment, but if the stage of disease is not coded in their primary care record^11^ they would not be automatically identified by NHS Digital; however, renal units may notify their stage 4 chronic kidney disease patients of their eligibility for treatment. In addition, NHS Digital holds only the General Practice Extraction Service (GPES) dataset, a substantially smaller derived subset of the full GP EHR dataset accessible through OpenSAFELY.

Where possible, we implemented the NHS Digital logic and associated codelists in the OpenSAFELY platform to identify patients in high risk groups. Codelists were used as published with the exception of minor adaptations made to i) code type where codes did not exist or were erroneous in the published codelist, and ii) code formatting for implementation in OpenSAFELY. Further details, including all limitations in implementing any of the logic or with the codelists are detailed in Supplementary material Table S1. If patients had records indicating that they fell into multiple high risk groups, all groups to which they belonged were used. The COVID-19 therapeutics dataset also included the high risk group(s) recorded by the clinician on the submitted form; if different or additional high risk groups were recorded here, patients were also assigned to these groups. Note that at the time of this analysis the clinician-assigned high risk group for patients receiving Paxlovid or Remdesivir was not available.

**Box 2:**
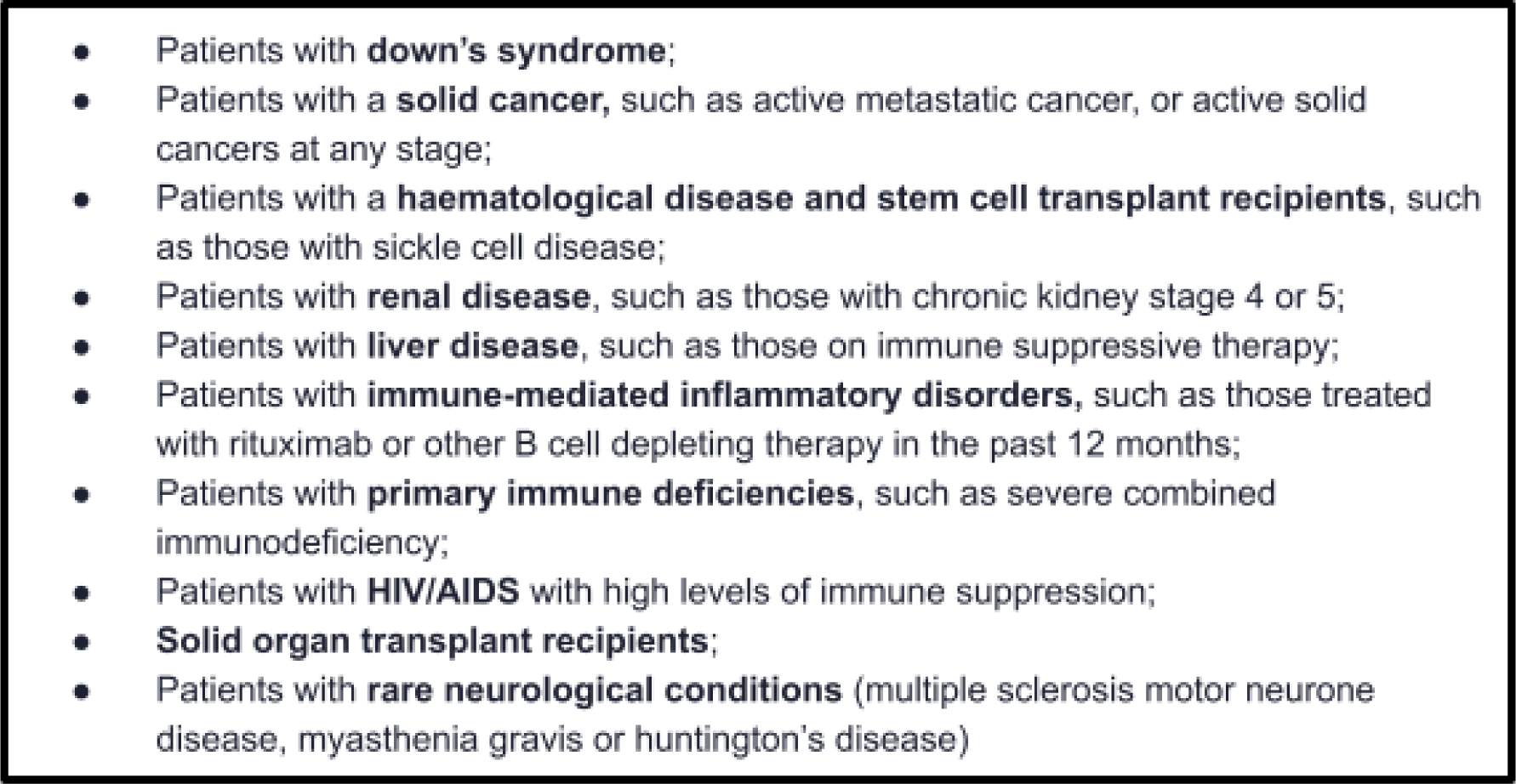
Patient groups considered at higher risk from COVID-19 and to be prioritised for treatment with antivirals and nMABs, as determined by an independent advisory group commissioned by the UK Department of Health and Social Care (DHSC)^7^. For further details on these criteria and how they were applied in the present study see Supplementary material Table S1

### Treated patients

We identified the subset of patients who received treatment in the COVID-19 therapeutics dataset, along with the treatment and the date they were given, restricted to those labelled “non_hospitalised”. We included first-line treatments Paxlovid and sotrovimab as per the most recent national policy at the time of this analysis^7^, as well as second- and third-line options remdesivir and molnupiravir respectively; sotrovimab and molnupiravir were available from the start of the study while Paxlovid and remdesivir were only available from 10^th^ February 2022. As previous versions of the policy also included casirivimab/imdevimab, patients who received this treatment were also included^12^.

### Key demographic and clinical characteristics of treated patients

We classified patients by age group, sex, NHS region of their general practice and other key demographics including ethnicity, the level of deprivation and rurality. Ethnicity was ascertained using 270 clinical codes grouped into broad categories; White, Black or Black British, Asian or Asian British, Mixed, Other, and Unknown. Deprivation was measured by Index of Multiple Deprivation (IMD), in quintiles, derived from the patient’s postcode at lower super output area level for a high degree of precision. Rurality was distinguished using the Rural Urban Classification^13^ and grouped into four broad categories; urban conurbation, urban city and town, rural town and fringe and rural - village and dispersed. Patients with missing sex, ethnicity, IMD, rurality or region were included as “Unknown”. Treated patients were also described according to whether they were in other groups of interest who are sometimes subject to variation in care^14^, including autism, dementia, learning disability, serious mental illness, care home residents, and housebound. Patients were also classified by their COVID-19 vaccination status (unvaccinated, unvaccinated with a record of declining vaccination, one vaccination, two vaccinations, or three or more).

### Consistency with guidance

For patients who received treatment but who were not otherwise identified as potentially being eligible for treatment, we report which eligibility or exclusion criteria were not met according to the data available (i.e. no positive SARS-CoV-2 test result, or not identified as a member of a high risk group). Where possible within available data, we also report other potential inconsistencies with guidance for patients who received treatment, such as where the high risk group identified within their records did not match the high risk group associated with their treatment.

We also assess consistency with treatment-specific criteria (as detailed in Table S2), such as patients having a recorded contraindication to the specific treatment given (e.g. adolescents treated with sotrovimab/remdesivir with weight under 40kg, Table S2), or patients treated outside the prescribed timescale, 5-7 days from symptom onset, depending on the treatment (Supplementary material Table S2). As symptom onset date was not available, here we used positive SARS-CoV-2 test as a proxy to estimate the extent to which patients may or may not have been treated outside the guidance time window.

### COVID Medicine Delivery Units

Details of all CMDUs can be found on the national website^15^. As CMDU identifiers were not available in OpenSafely, an alternative geographic grouping, Sustainability and Transformation Plan (STP), which has almost a 1:1 mapping, was used as a proxy to identify any variation in the proportion of patients treated between CMDUs. Note, the subset of the population covered by TPP in each STP may not be representative of the whole STP and STPs were only included if they had greater than 10% population coverage in TPP practices. Practice-STP mappings, used to calculate the coverage, were calculated as of March 2020 and it is likely that since then some borders and population sizes may have changed.

### Descriptive statistics

We generated charts showing the cumulative number of potentially eligible and treated patients per week, stratified by high risk group, and also stratified by treatment type for treated patients. We used simple descriptive statistics to summarise the counts and proportions of potentially eligible patients treated, stratified by treatment type and either high risk group or clinical and demographic groups, and to describe potential inconsistencies with guidelines.

### Codelists and implementation

Information on all covariates were obtained from primary care, secondary care and other records by searching TPP SystmOne records and linked datasets for specific coded data. Detailed information on compilation and sources for every individual codelist is available at https://www.opencodelists.org/ and all codelists are available under open licenses for review and re-use by the broader research community.

### Software and Reproducibility

All data were linked, stored and analysed securely through OpenSAFELY, a data analytics platform created by our team on behalf of NHS England to address urgent COVID-19 research questions (https://opensafely.org). All activity on the platform is publicly logged and all analytic code and supporting clinical codelists are automatically published at the time of results publication or sooner. In addition, the framework provides assurance that the analysis is reproducible and reusable. Further details on our information governance can be found on page 20, under information governance and ethics.

Data management and analysis was performed using the OpenSAFELY software libraries, Python 3 and R version 4.0.2. All code for the OpenSAFELY platform is freely available under open licenses for review and re-use on GitHub (https://github.com/opensafely). All code for data management and analysis for this paper is freely available under open licenses for review and re-use on GitHub (https://github.com/opensafely/antibody-and-antiviral-deployment).

### Reporting

This study followed STROBE-RECORD reporting guidelines. Charts and results not presented in this manuscript are available online for inspection in the associated GitHub repository^16^. To mitigate the risk of disclosure, patient counts of 0-7 are shown as “<8” with remaining counts rounded to the nearest 10 to protect against small number differences when routinely updating data. All percentages (%) are calculated with 95% confidence intervals (CI).

### Patient and Public Involvement

Patients were not formally involved in developing this specific study design, as it was developed in the context of the rapid rollout of a new treatment service during a global health emergency. We have developed a publicly available website https://opensafely.org/ through which we invite any patient or member of the public to contact us regarding this study or the broader OpenSAFELY project.

## RESULTS

### Eligibility for treatment

Between 11^th^ December 2021 and 28^th^ April 2022, a total of 102,170 patients registered at a TPP practice in England were identified as potentially being eligible for receiving an antiviral or nMAB for treating COVID-19 (4,690/102,170 patients were included due to having a record for receiving treatment, but whom were not otherwise considered eligible). The number of patients potentially eligible in each high risk group is described in Figure 1 and Table 1, with the greatest number of potentially eligible patients classified as those having an Immune mediated inflammatory disorder (n=40,360).

**Table 1.** Count and proportion of potentially eligible patients in OpenSAFELY-TPP who received treatment for COVID-19 between 16^th^ December 2021 and 28^th^ April 2022, broken down by high risk group and treatment type. Patient counts of 0-7 are shown as <8 with remaining counts rounded to the nearest 10; as a result percentages may not add up to 100%

| High risk cohort |  | Treated |  |  |  |  |  |  |  |  |  |  |  |
| --- | --- | --- | --- | --- | --- | --- | --- | --- | --- | --- | --- | --- | --- |
|  | Eligible | All |  | Paxlovid |  | Sotrovimab |  | Remdesivir |  | Molnupiravir |  | Casirivimab |  |
|  | Count | Count | % | Count | % | Count | % | Count | % | Count | % | Count | % |
| All | 102170 | 16930 | 17 (16-17) | 3060 | 18 (17-19) | 9290 | 55 (54-56) | 20 | 0 (0-0) | 4500 | 27 (26-27) | 50 | 0 (0-0) |
| Immune-mediated inflammatory disorders | 40360 | 7600 | 19 (18-19) | 1180 | 16 (15-16) | 4260 | 56 (55-57) | 10 | 0 (0-0) | 2130 | 28 (27-29) | 20 | 0 (0-0) |
| Primary immune deficiencies | 18910 | 3080 | 16 (16-17) | 730 | 24 (22-25) | 1520 | 49 (48-51) | <8 | -- | 820 | 27 (25-28) | 10 | 0 (0-1) |
| Solid cancer | 17840 | 2670 | 15 (14-15) | 350 | 13 (12-14) | 1580 | 59 (57-61) | <8 | -- | 730 | 27 (26-29) | 10 | 0 (0-1) |
| Rare neurological conditions | 11680 | 2620 | 22 (22-23) | 760 | 29 (27-31) | 1210 | 46 (44-48) | <8 | -- | 640 | 24 (23-26) | <8 | -- |
| Haematological diseases and stem cell transplant recipients | 9560 | 2620 | 27 (27-28) | 420 | 16 (15-17) | 1540 | 59 (57-61) | <8 | -- | 650 | 25 (23-26) | 10 | 0 (0-1) |
| Solid organ transplant recipients | 6690 | 1970 | 29 (28-31) | 50 | 3 (2-3) | 1400 | 71 (69-73) | <8 | -- | 500 | 25 (23-27) | <8 | -- |
| Renal disease | 6660 | 1930 | 29 (28-30) | <8 | -- | 1400 | 73 (71-75) | <8 | -- | 520 | 27 (25-29) | <8 | -- |
| Liver disease | 6240 | 750 | 12 (11-13) | 40 | 5 (4-7) | 480 | 64 (61-67) | <8 | -- | 240 | 32 (29-35) | <8 | -- |
| Down's syndrome | 2350 | 450 | 19 (18-21) | 130 | 29 (25-33) | 160 | 36 (31-40) | <8 | -- | 160 | 36 (31-40) | <8 | -- |
| Immunosuppression due to HIV or AIDS | 560 | 330 | 59 (55-63) | <8 | -- | 170 | 52 (46-57) | <8 | -- | 160 | 48 (43-54) | <8 | -- |

\* high risk groups are arranged in descending order, according to number of potentially eligible patients
† All percentages (%) are calculated with 95% confidence intervals

**Figure 1.**
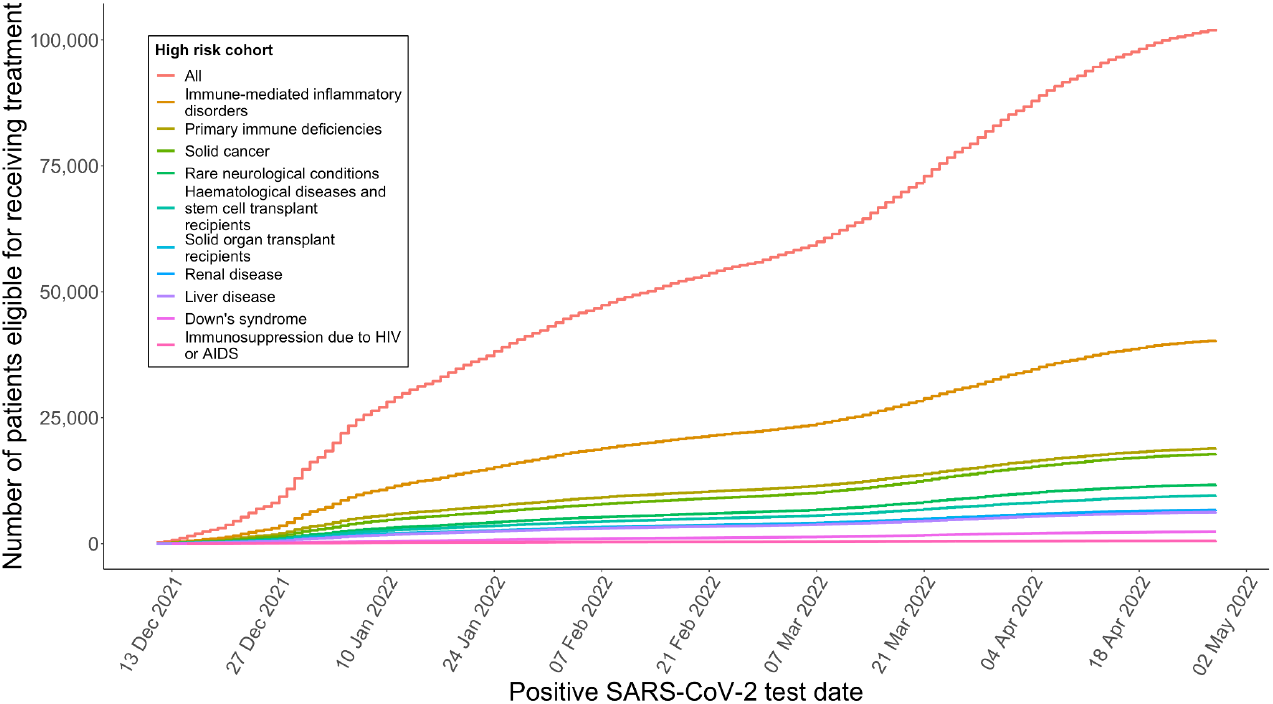
Cumulative total of potentially eligible patients for receiving an antiviral or nMABs for treating COVID-19 since 11^th^ December 2021, stratified by high risk group. Patients are considered eligible on the date of their positive SARS-CoV-2 test. Note, patients can appear in more than one high risk group, and the overall number in each group is likely to be an overestimation due to including SARS-CoV-2 infection confirmed by either lateral flow or PCR test (where only PCR-confirmed infections should have been treated according to guidance in effect prior to 10th February 2022), and potentially including non-symptomatic patients.

### Coverage of COVID-19 treatment

Of the 102,170 potentially eligible patients, 18,210 (18%) received treatment from a CMDU (Table 1; Figure 2; 220 patients were excluded due to having records of multiple treatments within a two week period, or an implausible treatment date). The proportion of potentially eligible patients receiving treatment increased over time, rising from 8% (180/2,380) in the first week of the roll out to 22% (420/1870) in the last week of April 2022 (Supplementary material Figure S1). Sotrovimab was the most widely used treatment over the study period (n=9,340, 51% of those treated) followed by molnupiravir (n=4,500, 25% of those treated) and Paxlovid (n=4,290, 24% of those treated). Use of casirivimab (n=50) and remdesivir (n=20) was low.

**Figure 2.**
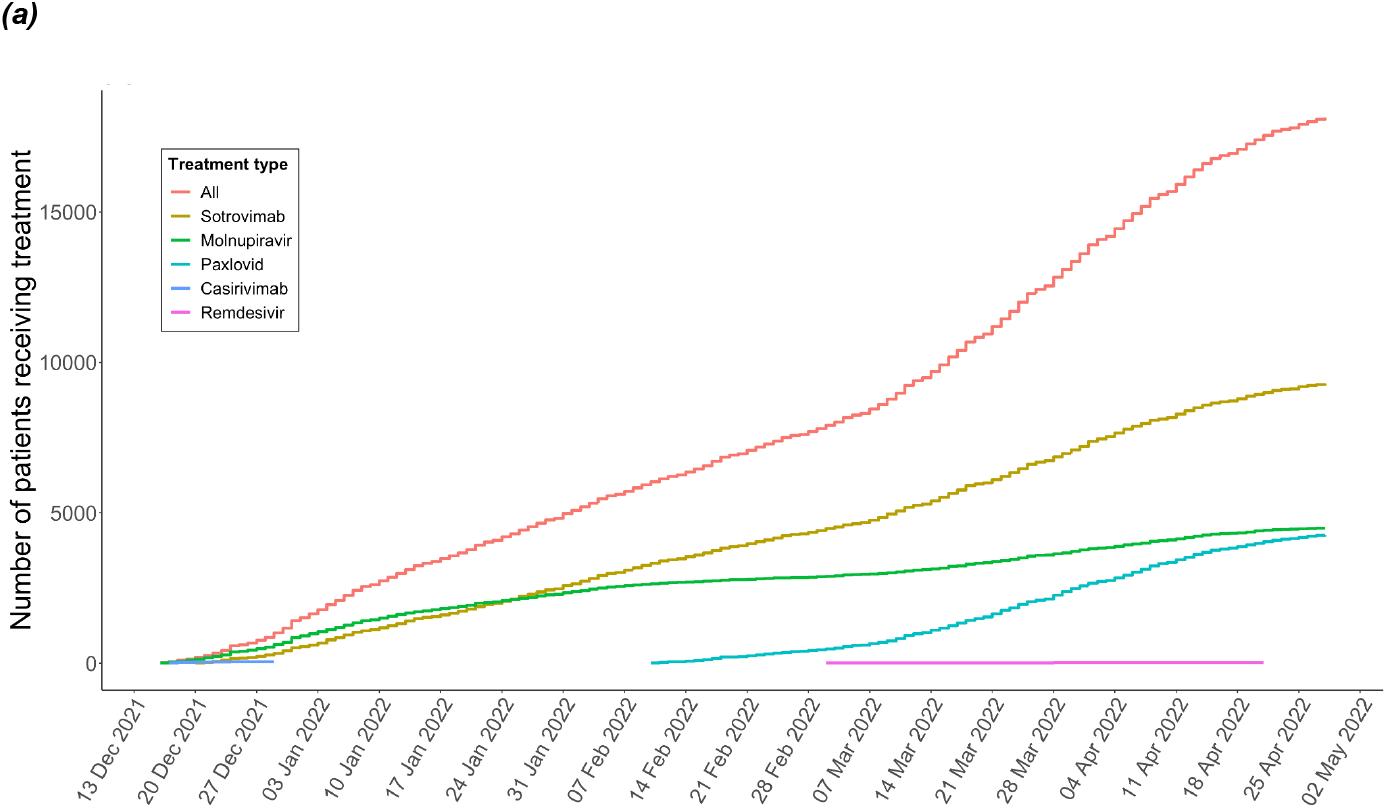
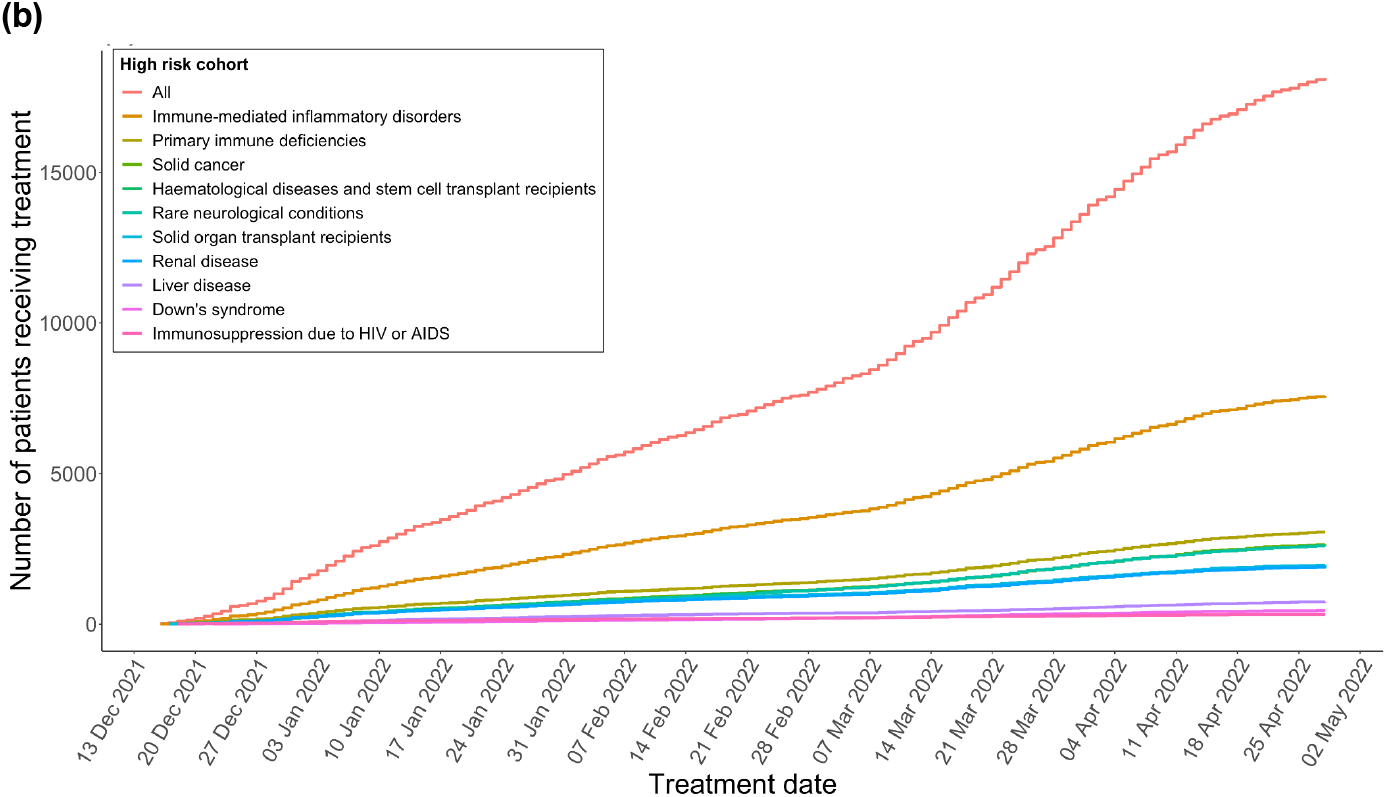
Cumulative total of patients in OpenSAFELY-TPP who received an antiviral or nMAB for treating COVID-19 since 16th December 2021, stratified by (a) treatment type and (b) high risk groups. Shorter lines for Paxlovid and casirivimab reflect availability and guidance. Note, patients can appear in more than one high risk group.

A sensitivity analysis restricting the population to only those with a “symptomatic” flag associated with their positive SARS-CoV-2 test reduced the potentially eligible population from 102,170 to 23,380 and the treated population from 18,210 to 4,500; a 77% and 75% reduction, respectively, and equivalent to 19% of patients receiving treatment from a CMDU.

### High risk patient groups

Of the potentially eligible patients, 96% (n=98,170) were assigned to at least one EHR-derived high risk group with an additional 3% (n=2,720) assigned to at least one clinician-assigned high risk cohort(s) in the COVID-19 therapeutics dataset. Of the 18,210 patients who received treatment, 78% (n=14,210) were assigned to at least one EHR-derived high risk group, of whom almost all (98%, n=13,950) matched the clinician-assigned high risk cohort(s). The majority (83%, n=3,320) of the 22% (n=4,000) who were treated but did not have an EHR-derived high risk cohort had a clinician-assigned high risk group of Immune-mediated inflammatory disorder, were treated with Paxlovid or remdesivir (for which the clinician-assigned high risk group was not available) or had a clinician-assigned high risk group of Solid cancer; 33% (n=1,300), 32% (n=1,280) and 19% (n=740), respectively.

The proportion of potentially eligible patients receiving treatment varied by high risk group (Table 1). For example: 19% (95% CI 18 to 19) of patients in the largest eligible group, Immune-mediated inflammatory disorders, received treatment; values ranged from 12% (95% CI 11 to 13) in Liver disease, to 29% (95% CI 28 to 31) in Solid organ transplant recipients. The type of treatment given also varied by high risk group with sotrovimab favoured over molnupiravir or Paxlovid in all but three high risk groups: Down syndrome (36%; 95% CI 31 to 40), Rare neurological conditions (46%; 95% CI 44 to 48), and Primary immune deficiencies (49%; 95% CI 48 to 51).

### Key demographic and clinical characteristics of treated patients

Table 2 shows the count and proportion of potentially eligible patients who received treatment for COVID-19 by the 28^th^ April 2022, broken down by demographic and clinical categories and by treatment type. Among eligible patients who received treatment, patients were more likely to be White (18%; 95% CI 18 to 19) or Asian or Asian British (15%; 95% CI 14 to 16) than Black or Black British (10%; 95% CI 9 to 11). The percentage treated correlated with deprivation quintile with 12% (95% CI 12 to 13) receiving treatment in the most deprived areas, compared to 21% (95% CI 21 to 22) in the least deprived areas, and similarly with rurality with 14% (95% CI 13 to 14) in “Urban - conurbation” compared to 22% (95% CI 21 to 23) in “Rural - village and dispersed”. The region with the highest rate of treatment was the East of England (23%; 95% CI 22 to 24) and the lowest was Yorkshire and the Humber (11%; 95% CI 10 to 12). Some clinical groups were much less likely to be treated; dementia (5%; 95% CI 4 to 6), care home residents (6%; 95% CI 5 to 6), and others had a slightly reduced chance; patients with sickle cell disease (10%; 95% CI 8 to 11), those who were housebound (13%; 95% CI 12 to 14), or those who had a severe mental illness (13%; 95% CI 11 to 15). Those classed as clinically extremely vulnerable were slightly more likely to be treated (22%; 95% CI 22 to 23). Unvaccinated patients were substantially less likely to receive treatment (5%; 95% CI 4 to 7).

**Table 2:**
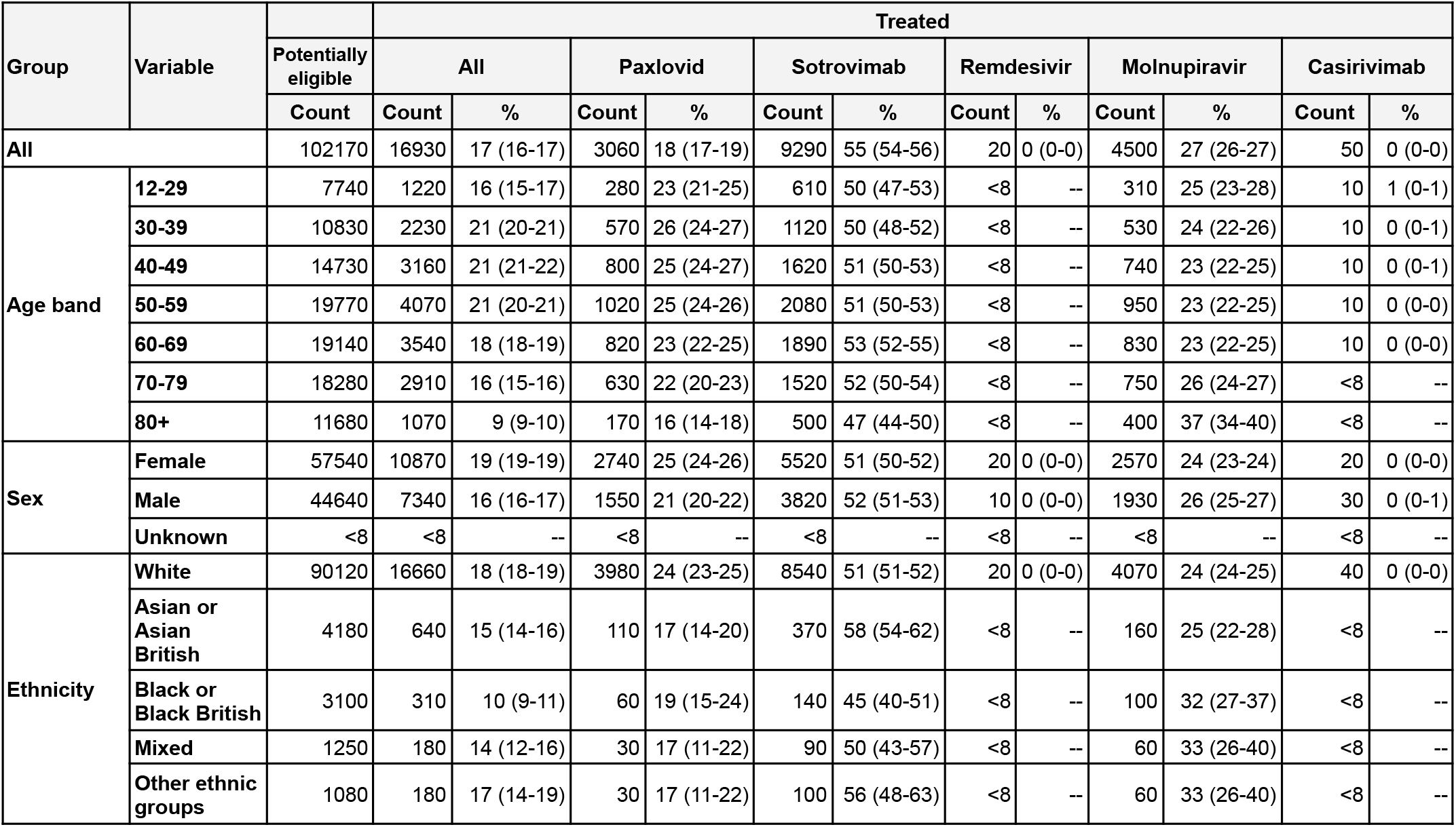
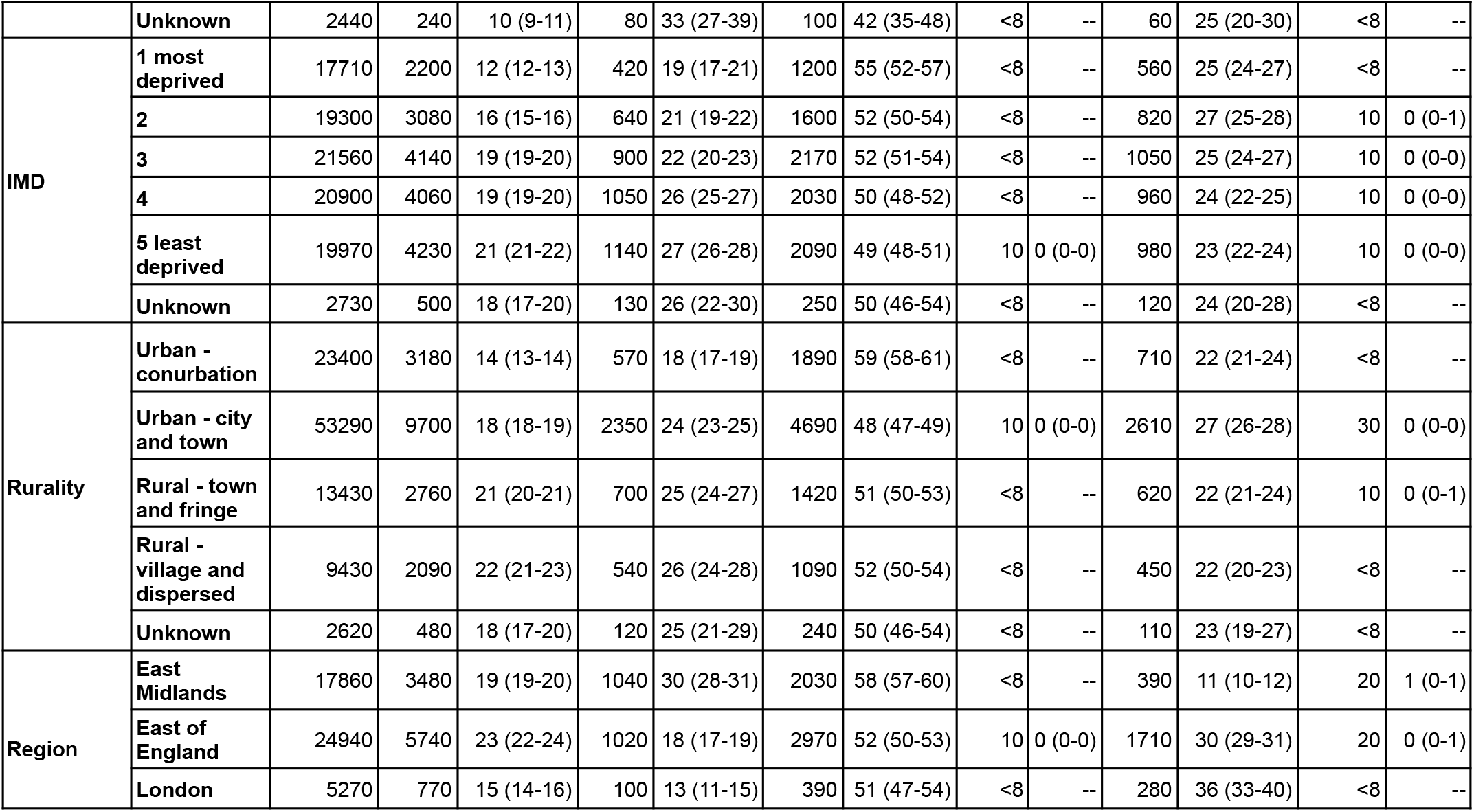
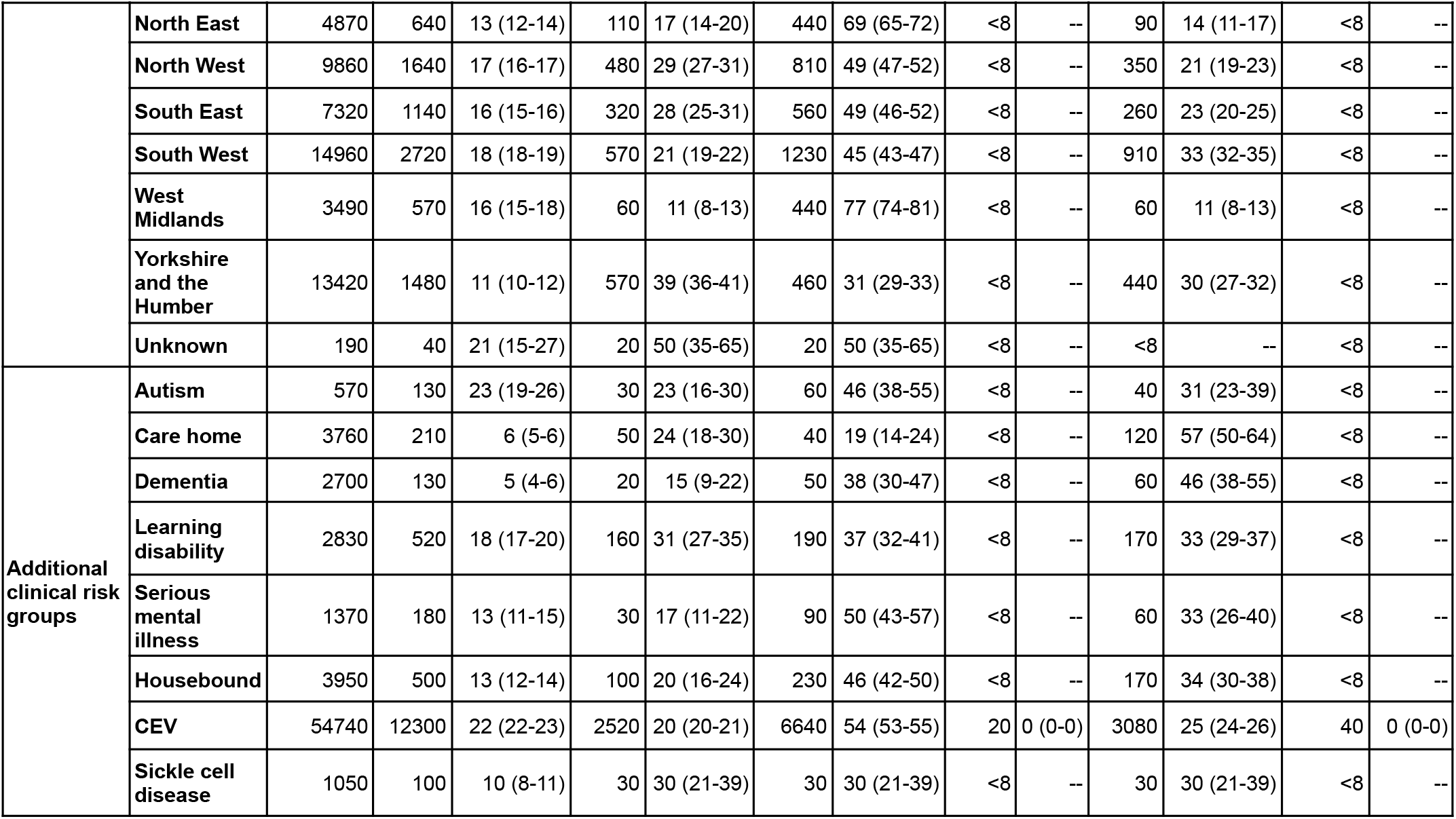
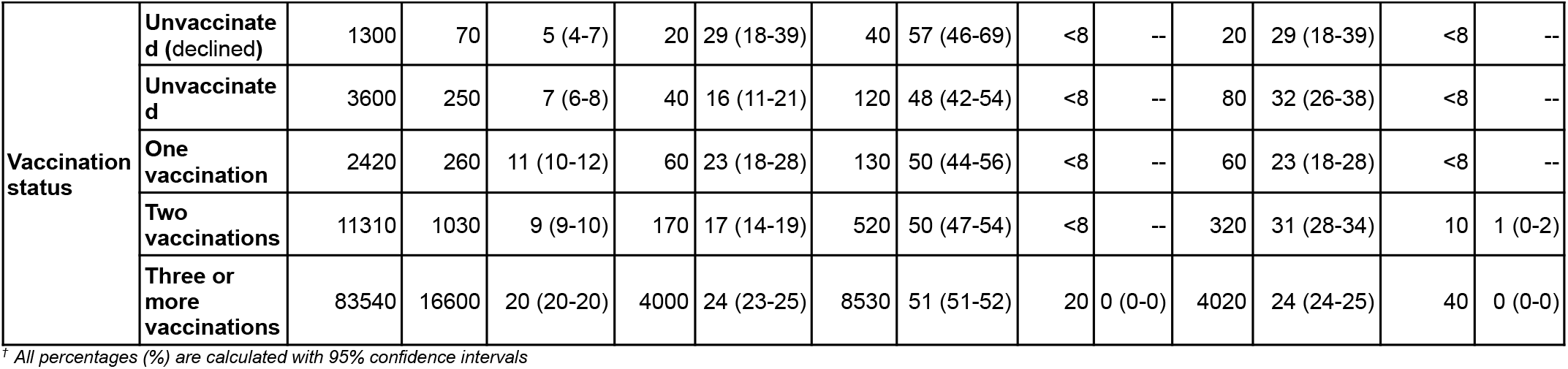
Count and proportion of potentially eligible patients in OpenSAFELY-TPP who received treatment for COVID-19 between 11th December 2021 and 28^th^ April 2022, broken down by demographic and clinical categories and by treatment type. Patient counts of 0-7 are shown as <8 with remaining counts rounded to the nearest 10; as a result percentages may not add up to 100%

| Group | Variable |  | Treated |  |  |  |  |  |  |  |  |  |  |  |
| --- | --- | --- | --- | --- | --- | --- | --- | --- | --- | --- | --- | --- | --- | --- |
|  |  | Potentially eligible | All |  | Paxlovid |  | Sotrovimab |  | Remdesivir |  | Molnupiravir |  | Casirivimab |  |
|  |  | Count | Count | % | Count | % | Count | % | Count | % | Count | % | Count | % |
| All |  | 102170 | 16930 | 17 (16-17) | 3060 | 18 (17-19) | 9290 | 55 (54-56) | 20 | 0 (0-0) | 4500 | 27 (26-27) | 50 | 0 (0-0) |
| Age band | 12-29 | 7740 | 1220 | 16 (15-17) | 280 | 23 (21-25) | 610 | 50 (47-53) | <8 | -- | 310 | 25 (23-28) | 10 | 1 (0-1) |
|  | 30-39 | 10830 | 2230 | 21 (20-21) | 570 | 26 (24-27) | 1120 | 50 (48-52) | <8 | -- | 530 | 24 (22-26) | 10 | 0 (0-1) |
|  | 40-49 | 14730 | 3160 | 21 (21-22) | 800 | 25 (24-27) | 1620 | 51 (50-53) | <8 | -- | 740 | 23 (22-25) | 10 | 0 (0-1) |
|  | 50-59 | 19770 | 4070 | 21 (20-21) | 1020 | 25 (24-26) | 2080 | 51 (50-53) | <8 | -- | 950 | 23 (22-25) | 10 | 0 (0-0) |
|  | 60-69 | 19140 | 3540 | 18 (18-19) | 820 | 23 (22-25) | 1890 | 53 (52-55) | <8 | -- | 830 | 23 (22-25) | 10 | 0 (0-0) |
|  | 70-79 | 18280 | 2910 | 16 (15-16) | 630 | 22 (20-23) | 1520 | 52 (50-54) | <8 | -- | 750 | 26 (24-27) | <8 | -- |
|  | 80+ | 11680 | 1070 | 9 (9-10) | 170 | 16 (14-18) | 500 | 47 (44-50) | <8 | -- | 400 | 37 (34-40) | <8 | -- |
| Sex | Female | 57540 | 10870 | 19 (19-19) | 2740 | 25 (24-26) | 5520 | 51 (50-52) | 20 | 0 (0-0) | 2570 | 24 (23-24) | 20 | 0 (0-0) |
|  | Male | 44640 | 7340 | 16 (16-17) | 1550 | 21 (20-22) | 3820 | 52 (51-53) | 10 | 0 (0-0) | 1930 | 26 (25-27) | 30 | 0 (0-1) |
|  | Unknown | <8 | <8 | -- | <8 | -- | <8 | -- | <8 | -- | <8 | -- | <8 | -- |
| Ethnicity | White | 90120 | 16660 | 18 (18-19) | 3980 | 24 (23-25) | 8540 | 51 (51-52) | 20 | 0 (0-0) | 4070 | 24 (24-25) | 40 | 0 (0-0) |
|  | Asian or Asian British | 4180 | 640 | 15 (14-16) | 110 | 17 (14-20) | 370 | 58 (54-62) | <8 | -- | 160 | 25 (22-28) | <8 | -- |
|  | Black or Black British | 3100 | 310 | 10 (9-11) | 60 | 19 (15-24) | 140 | 45 (40-51) | <8 | -- | 100 | 32 (27-37) | <8 | -- |
|  | Mixed | 1250 | 180 | 14 (12-16) | 30 | 17 (11-22) | 90 | 50 (43-57) | <8 | -- | 60 | 33 (26-40) | <8 | -- |
|  | Other ethnic groups | 1080 | 180 | 17 (14-19) | 30 | 17 (11-22) | 100 | 56 (48-63) | <8 | -- | 60 | 33 (26-40) | <8 | -- |
|  |  | Potentially eligible | All |  | Paxlovid |  | Sotrovimab |  | Remdesivir |  | Molnupiravir |  | Casirivimab |  |
|  |  | Count | Count | % | Count | % | Count | % | Count | % | Count | % | Count | % |
|  | Unknown | 2440 | 240 | 10 (9-11) | 80 | 33 (27-39) | 100 | 42 (35-48) | <8 | -- | 60 | 25 (20-30) | <8 | -- |
| IMD | 1 most deprived | 17710 | 2200 | 12 (12-13) | 420 | 19 (17-21) | 1200 | 55 (52-57) | <8 | -- | 560 | 25 (24-27) | <8 | -- |
|  | 2 | 19300 | 3080 | 16 (15-16) | 640 | 21 (19-22) | 1600 | 52 (50-54) | <8 | -- | 820 | 27 (25-28) | 10 | 0 (0-1) |
|  | 3 | 21560 | 4140 | 19 (19-20) | 900 | 22 (20-23) | 2170 | 52 (51-54) | <8 | -- | 1050 | 25 (24-27) | 10 | 0 (0-0) |
|  | 4 | 20900 | 4060 | 19 (19-20) | 1050 | 26 (25-27) | 2030 | 50 (48-52) | <8 | -- | 960 | 24 (22-25) | 10 | 0 (0-0) |
|  | 5 least deprived | 19970 | 4230 | 21 (21-22) | 1140 | 27 (26-28) | 2090 | 49 (48-51) | 10 | 0 (0-0) | 980 | 23 (22-24) | 10 | 0 (0-0) |
|  | Unknown | 2730 | 500 | 18 (17-20) | 130 | 26 (22-30) | 250 | 50 (46-54) | <8 | -- | 120 | 24 (20-28) | <8 | -- |
| Rurality | Urban - conurbation | 23400 | 3180 | 14 (13-14) | 570 | 18 (17-19) | 1890 | 59 (58-61) | <8 | -- | 710 | 22 (21-24) | <8 | -- |
|  | Urban - city and town | 53290 | 9700 | 18 (18-19) | 2350 | 24 (23-25) | 4690 | 48 (47-49) | 10 | 0 (0-0) | 2610 | 27 (26-28) | 30 | 0 (0-0) |
|  | Rural - town and fringe | 13430 | 2760 | 21 (20-21) | 700 | 25 (24-27) | 1420 | 51 (50-53) | <8 | -- | 620 | 22 (21-24) | 10 | 0 (0-1) |
|  | Rural - village and dispersed | 9430 | 2090 | 22 (21-23) | 540 | 26 (24-28) | 1090 | 52 (50-54) | <8 | -- | 450 | 22 (20-23) | <8 | -- |
|  | Unknown | 2620 | 480 | 18 (17-20) | 120 | 25 (21-29) | 240 | 50 (46-54) | <8 | -- | 110 | 23 (19-27) | <8 | -- |
| Region | East Midlands | 17860 | 3480 | 19 (19-20) | 1040 | 30 (28-31) | 2030 | 58 (57-60) | <8 | -- | 390 | 11 (10-12) | 20 | 1 (0-1) |
|  | East of England | 24940 | 5740 | 23 (22-24) | 1020 | 18 (17-19) | 2970 | 52 (50-53) | 10 | 0 (0-0) | 1710 | 30 (29-31) | 20 | 0 (0-1) |
|  | London | 5270 | 770 | 15 (14-16) | 100 | 13 (11-15) | 390 | 51 (47-54) | <8 | -- | 280 | 36 (33-40) | <8 | -- |

| Group | Variable | Potentially eligible | Treated |  |  |  |  |  |  |  |  |  |  |  |
| --- | --- | --- | --- | --- | --- | --- | --- | --- | --- | --- | --- | --- | --- | --- |
|  |  |  | All |  | Paxlovid |  | Sotrovimab |  | Remdesivir |  | Molnupiravir |  | Casirivimab |  |
|  |  |  | Count | % | Count | % | Count | % | Count | % | Count | % | Count | % |
|  | North East | 4870 | 640 | 13 (12-14) | 110 | 17 (14-20) | 440 | 69 (65-72) | <8 | -- | 90 | 14 (11-17) | <8 | -- |
|  | North West | 9860 | 1640 | 17 (16-17) | 480 | 29 (27-31) | 810 | 49 (47-52) | <8 | -- | 350 | 21 (19-23) | <8 | -- |
|  | South East | 7320 | 1140 | 16 (15-16) | 320 | 28 (25-31) | 560 | 49 (46-52) | <8 | -- | 260 | 23 (20-25) | <8 | -- |
|  | South West | 14960 | 2720 | 18 (18-19) | 570 | 21 (19-22) | 1230 | 45 (43-47) | <8 | -- | 910 | 33 (32-35) | <8 | -- |
|  | West Midlands | 3490 | 570 | 16 (15-18) | 60 | 11 (8-13) | 440 | 77 (74-81) | <8 | -- | 60 | 11 (8-13) | <8 | -- |
|  | Yorkshire and the Humber | 13420 | 1480 | 11 (10-12) | 570 | 39 (36-41) | 460 | 31 (29-33) | <8 | -- | 440 | 30 (27-32) | <8 | -- |
|  | Unknown | 190 | 40 | 21 (15-27) | 20 | 50 (35-65) | 20 | 50 (35-65) | <8 | -- | <8 | -- | <8 | -- |
| Additional clinical risk groups | Autism | 570 | 130 | 23 (19-26) | 30 | 23 (16-30) | 60 | 46 (38-55) | <8 | -- | 40 | 31 (23-39) | <8 | -- |
|  | Care home | 3760 | 210 | 6 (5-6) | 50 | 24 (18-30) | 40 | 19 (14-24) | <8 | -- | 120 | 57 (50-64) | <8 | -- |
|  | Dementia | 2700 | 130 | 5 (4-6) | 20 | 15 (9-22) | 50 | 38 (30-47) | <8 | -- | 60 | 46 (38-55) | <8 | -- |
|  | Learning disability | 2830 | 520 | 18 (17-20) | 160 | 31 (27-35) | 190 | 37 (32-41) | <8 | -- | 170 | 33 (29-37) | <8 | -- |
|  | Serious mental illness | 1370 | 180 | 13 (11-15) | 30 | 17 (11-22) | 90 | 50 (43-57) | <8 | -- | 60 | 33 (26-40) | <8 | -- |
|  | Housebound | 3950 | 500 | 13 (12-14) | 100 | 20 (16-24) | 230 | 46 (42-50) | <8 | -- | 170 | 34 (30-38) | <8 | -- |
|  | CEV | 54740 | 12300 | 22 (22-23) | 2520 | 20 (20-21) | 6640 | 54 (53-55) | 20 | 0 (0-0) | 3080 | 25 (24-26) | 40 | 0 (0-0) |
|  | Sickle cell disease | 1050 | 100 | 10 (8-11) | 30 | 30 (21-39) | 30 | 30 (21-39) | <8 | -- | 30 | 30 (21-39) | <8 | -- |
|  |  | Potentially eligible | All |  | Paxlovid |  | Sotrovimab |  | Remdesivir |  | Molnupiravir |  | Casirivimab |  |
|  |  | Count | Count | % | Count | % | Count | % | Count | % | Count | % | Count | % |
| Vaccination status | Unvaccinated (declined) | 1300 | 70 | 5 (4-7) | 20 | 29 (18-39) | 40 | 57 (46-69) | <8 | -- | 20 | 29 (18-39) | <8 | -- |
|  | Unvaccinated | 3600 | 250 | 7 (6-8) | 40 | 16 (11-21) | 120 | 48 (42-54) | <8 | -- | 80 | 32 (26-38) | <8 | -- |
|  | One vaccination | 2420 | 260 | 11 (10-12) | 60 | 23 (18-28) | 130 | 50 (44-56) | <8 | -- | 60 | 23 (18-28) | <8 | -- |
|  | Two vaccinations | 11310 | 1030 | 9 (9-10) | 170 | 17 (14-19) | 520 | 50 (47-54) | <8 | -- | 320 | 31 (28-34) | 10 | 1 (0-2) |
|  | Three or more vaccinations | 83540 | 16600 | 20 (20-20) | 4000 | 24 (23-25) | 8530 | 51 (51-52) | 20 | 0 (0-0) | 4020 | 24 (24-25) | 40 | 0 (0-0) |

<sup>†</sup> All percentages (%) are calculated with 95% confidence intervals

### Consistency with guidance

Of the 18,210 patients who received treatment for COVID-19, 6% (n=1,140) did not have evidence of a positive SARS-CoV-2 test, 22% (n=4,000) did not have an EHR-derived high risk group, and <1% (n=30) were discharged from hospital within 30 days prior to positive test or treatment, where COVID-19 was the primary diagnosis. There were a small number of other potential inconsistencies with guidance for patients who received treatment (Supplementary material Figure S2), such as a potential contraindication: of the contraindications included, the only one identified in results was recorded adolescent weight ≤40kg for sotrovimab (<1%; n=10) (see Discussion).

Overall, of patients who received treatment, 96% (n=16,370) did so within the respective treatment-specific eligibility window as estimated from their positive SARS-CoV-2 test date (as symptom onset date was not consistently available) (Supplementary material Figures S3 and S4). Treatment occurred most commonly two days (33%; n=5,640) after a patient’s positive SARS-CoV-2 test. There was minor variation between the three most common treatments: treatment with Paxlovid occurred slightly earlier compared to molnupiravir and sotrovimab with 76% (n=2,990) versus 63% (n=2,640) and 57% (n=4,970) of patients being treated within two days of their positive test, respectively.

### COVID Medicine Delivery Units

STP (used as a proxy for CMDU) was available for all eligible patients, with a total of 26 STPs identified and included (six were excluded due to having less than 10% population coverage in TPP practices; 2,650 eligible patients and 220 treated patients). The overall proportion of potentially eligible patients receiving treatment varied by STP (Supplementary material Figure S5) from 11% (n=8,910) in STPs in the lowest decile, to 25% (n=12,210) in STPs in the highest decile. In addition, the maximum weekly proportion of patients treated over the study period ranged from 17% in STPs in the lowest decile, to 39% in STPs in the highest decile.

## Discussion

### Summary

The NHS in England rapidly established CMDUs to support delivery of COVID-19 therapeutics to people in the community from December 2021. In our study, out of 102,170 patients likely to be eligible based on national clinical criteria, 18,210 (18%) received an antiviral or nMAB between 16^th^ December 2021 and 28^th^ April 2022. Sotrovimab was the most widely used treatment, followed by molnupiravir and Paxlovid (51%, 25% and 24%, respectively), although use varied over time. There was limited use of other treatments, reflective of availability and guidance. The proportion of the potentially eligible patients receiving treatment increased over time, rising from 8% in the first week of treatment availability to 22% in the latest week, and varied by high risk group, e.g. 29% in Solid organ transplant recipients, 12% in Liver disease. Treatment type also varied by high risk group, with sotrovimab favoured over molnupiravir/Paxlovid in all but three high risk groups: Down syndrome (36%), Rare neurological conditions (46%), and Primary immune deficiencies (49%). We observed differences in treatment rates between demographic and clinical sub-groups. Those living in more socioeconomically deprived areas generally had lower treatment coverage (12% in the most deprived quintile, vs: 21% in the least deprived quintile), as did those living in care homes or who were housebound (6% and 13%, respectively). Patients who were White or Asian or Asian British were most likely to receive treatment, with Black or Black British patients the least likely (18% and 15% vs 10%, respectively). We observed substantial geographic variation in treatment rates between NHS Regions (from 11% in Yorkshire and the Humber to 23% in the East of England) and between STPs (11% in STPs in the lowest decile, to 25% in STPs in the highest decile).

### Strengths and weaknesses

The key strengths of this study are the scale, detail and completeness of the underlying raw EHR data. The OpenSAFELY-TPP platform runs analyses across the full dataset of all raw, pseudonymised, single-event-level clinical events for all 23.4 million current patients at all 2,545 general practices in England using TPP software; whereas the GPES dataset available in NHS Digital is a subset of this raw data created through a series of processing rules for specific aspects of GP records applied at source before extraction. OpenSAFELY-TPP also provides data in near-real time, providing unprecedented opportunities for audit and feedback to rapidly identify and resolve concerns around health service activity and clinical outcomes related to the COVID-19 pandemic. The delay from entry of a clinical event into the EHR to its appearing in the OpenSAFELY-TPP platform varies from two to nine days. This is substantially faster than any other source of comprehensive GP data. Additionally OpenSAFELY now contains linked COVID-19 therapeutics data which is collected from CMDUs with typically only 2-3 days delay between form submission and data being available for analysis. This can support timely monitoring of treatments administered through CMDUs as well as safety and efficacy studies of these treatments and other COVID-19 analyses.

We recognise some limitations to our analysis. Our population, although extremely large, may not be fully representative: there is some geographic clustering in the EHR system used by general practices, and only 17% of general practices in London use TPP software. However other GP data sources such as the CPRD data download service similarly contain a convenience sample (practices that have elected to participate in data extraction) rather than a random sample of the general population; and there are no a priori reasons to expect that this issue will substantially affect the relationships observed between patient factors and outcomes.

As with other analyses of the same data, the newly sourced COVID-19 therapeutics data represent treatment notifications, not prescriptions, and as such there may be some missingness caused by delays in paperwork being completed. In addition, this data does not allow us to determine the reasons why some patients who were considered eligible may not have received treatment; many patients would not have been treated because they were asymptomatic or clinically improving which can not be elucidated from the data. Because the presence of symptoms could not be reliably determined for the purposes of inclusion criteria (as highlighted by our sensitivity analysis which showed that restricting to only those with a “symptomatic” flag associated with their positive SARS-CoV-2 test reduced the population by over 70%) and as it was not possible distinguish between PCR and lateral flow tests, we are likely to somewhat overestimate the total number of potentially eligible people.

EHR data may not always fully capture some eligibility criteria and as such, may underestimate the true number of eligible people in some groups or misclassify some people, particularly those identified through “non-digital” routes, e.g. patients with kidney disease. Related to this, as previously described, we may not have ascertained all people in the Immunosuppression due to HIV/AIDS group due to specific arrangements around HIV data^17^. In addition, our ascertainment of eligibility status may sometimes deviate from NHS Digital ascertained eligibility status on specific patients for two reasons: OpenSAFELY has different and more detailed primary care records available; and when translating information from the NHS Digital website into analytic code, we had to make pragmatic decisions to resolve some discrepancies (as described). We have notified NHS Digital of discrepancies we identified with their codelists - which we regard as normal and expected with complex cohorting work - and additionally made all our analytic code and codelists openly available for inspection, as with all OpenSAFELY analyses. It is additionally reasonable to expect marginal differences in the proportion of the potentially eligible in each risk group who were treated between different data sources from different analytic teams due to minor differences in the speeds of data flow, specific data available, and the ascertainment of denominators. Finally, our findings on apparent inconsistencies with treatment guidance should be taken as indicative only, as there were several limitations such as: the date of treatment may occasionally be entered incorrectly on the submitted form; some SARS-CoV-2 test records may not pass into EHRs; hospitalisation due to COVID-19 is difficult to determine accurately in HES data (as it is not possible to fully determine whether the patient was treated for COVID-19 during hospitalisation or if it was an incidental finding); and the latest weight recorded in EHR (required to be over 40kg for adolescents treated with sotrovimab or remdesivir) may not be current at the time of treatment.

### Policy Implications and future research

To our knowledge this paper is the first study to describe in detail the demographic and clinical features of those who received treatments from CMDUs across England; and the first to report variation in treatment by detailed demographic and clinical characteristics. Our finding that only 18% of potentially eligible patients received treatment could be of concern, however, this may just reflect the number of patients who were asymptomatic or improving by the time they were assessed by the CMDU. While these clinical data are not captured in the primary care record or the CMDU data, we understand that CMDU data recording is being iterated to capture some reasons for non-treatment among those who were assessed; we will update our analyses as this data becomes available. Although sotrovimab was the first-line treatment available during the majority of the study period, almost 50% of patients received molnupiravir or Paxlovid; this may be explained by sotrovimab requiring an infusion in a clinic setting, presenting greater logistical challenges. Our findings of discrepancies in treatment between different groups is notable and consistent with our analysis of sociodemographic and ethnic variation in the receipt of COVID-19 vaccinations^8^: Further research and investigation is required to understand and address the causes of any inequity. Additionally, understanding variation in the choice of individual treatment option beyond clinical indications and cautions will be of utmost importance as more information on the relative efficacy emerges from ongoing trials.

The reasons underpinning variation in treatments delivered by CMDUs are not yet understood, and information presented here should not be misinterpreted as a criticism of the rapidly established CMDUs in the context of high levels of infection^18^, but rather as an example of the value of rapid turnaround data monitoring to help optimise the successful delivery of an ambitious national treatment programme. We will produce routine data updates at https://reports.opensafely.org/ to assist with ongoing monitoring and targeted initiatives to address gaps in coverage as well as informing development of study designs on the efficacy and safety of these new treatments. It should lastly be noted that all findings in this paper are from the first few preliminary weeks of these treatments being made nationally available: substantial changes in coverage among different groups are to be expected over the coming months.

### Conclusions

The NHS in England has rapidly deployed facilities to offer novel therapeutics for the rapid treatment of COVID-19 in the community. Targeted activity may be needed to address lower treatment rates observed among certain geographic areas and key groups including ethnic minorities, people living in areas of higher deprivation, and in care homes. Near real-time data monitoring can help support those on the front line making complex operational decisions around treatment delivery.

## Data Availability

Access to the underlying identifiable and potentially re-identifiable pseudonymised electronic health record data is tightly governed by various legislative and regulatory frameworks, and restricted by best practice. The data in OpenSAFELY is drawn from General Practice data across England where TPP is the Data Processor. TPP developers (CB, JC, and SH) initiate an automated process to create pseudonymised records in the core OpenSAFELY database, which are copies of key structured data tables in the identifiable records. These are linked onto key external data resources that have also been pseudonymised via SHA-512 one-way hashing of NHS numbers using a shared salt. DataLab developers and PIs holding contracts with NHS England have access to the OpenSAFELY pseudonymised data tables as needed to develop the OpenSAFELY tools. These tools in turn enable researchers with OpenSAFELY Data Access Agreements to write and execute code for data management and data analysis without direct access to the underlying raw pseudonymised patient data, and to review the outputs of this code. All code for the full data management pipeline, from raw data to completed results for this analysis, and for the OpenSAFELY platform as a whole is available for review at github.com/OpenSAFELY.

https://github.com/opensafely/antibody-and-antiviral-deployment

## Abbreviations

(CMDUs): COVID-19 Medicines Units
(nMABs): neutralising monoclonal antibodies

## Administrative

## Acknowledgements

We are very grateful for all the support received from the EMIS and TPP Technical Operations team throughout this work, and for generous assistance from the information governance and database teams at NHS England / NHSX.

## Information governance and ethical approval

NHS England is the data controller; TPP is the data processor; and the researchers on OpenSAFELY are acting with the approval of NHS England. This implementation of OpenSAFELY is hosted within the TPP environment which is accredited to the ISO 27001 information security standard and is NHS IG Toolkit compliant;^19,20^ patient data has been pseudonymised for analysis and linkage using industry standard cryptographic hashing techniques; all pseudonymised datasets transmitted for linkage onto OpenSAFELY are encrypted; access to the platform is via a virtual private network (VPN) connection, restricted to a small group of researchers; the researchers hold contracts with NHS England and only access the platform to initiate database queries and statistical models; all database activity is logged; only aggregate statistical outputs leave the platform environment following best practice for anonymisation of results such as statistical disclosure control for low cell counts.^21^ The OpenSAFELY research platform adheres to the obligations of the UK General Data Protection Regulation (GDPR) and the Data Protection Act 2018. In March 2020, the Secretary of State for Health and Social Care used powers under the UK Health Service (Control of Patient Information) Regulations 2002 (COPI) to require organisations to process confidential patient information for the purposes of protecting public health, providing healthcare services to the public and monitoring and managing the COVID-19 outbreak and incidents of exposure; this sets aside the requirement for patient consent.^22^ Taken together, these provide the legal bases to link patient datasets on the OpenSAFELY platform. GP practices, from which the primary care data are obtained, are required to share relevant health information to support the public health response to the pandemic, and have been informed of the OpenSAFELY analytics platform.

This study was approved by the Health Research Authority (REC reference 20/LO/0651) and by the LSHTM Ethics Board (reference 21863).

## Data access and verification

Access to the underlying identifiable and potentially re-identifiable pseudonymised electronic health record data is tightly governed by various legislative and regulatory frameworks, and restricted by best practice. The data in OpenSAFELY is drawn from General Practice data across England where TPP is the Data Processor. TPP developers (CB, JC, and SH) initiate an automated process to create pseudonymised records in the core OpenSAFELY database, which are copies of key structured data tables in the identifiable records. These are linked onto key external data resources that have also been pseudonymised via SHA-512 one-way hashing of NHS numbers using a shared salt. DataLab developers and PIs (BG,) holding contracts with NHS England have access to the OpenSAFELY pseudonymised data tables as needed to develop the OpenSAFELY tools. These tools in turn enable researchers with OpenSAFELY Data Access Agreements to write and execute code for data management and data analysis without direct access to the underlying raw pseudonymised patient data, and to review the outputs of this code. All code for the full data management pipeline—from raw data to completed results for this analysis—and for the OpenSAFELY platform as a whole is available for review at github.com/OpenSAFELY.

## Authors’ contributions

Contributions are as follows:

Conceptualisation: AG, HC, SE, LT, BMK, BG

Funding acquisition: BG

Methodology: AG, HC, SE, LT, BMK, BG

Formal analysis: AG, HC

Codelists: RH, PI, AG, BMK

Software: RS, SB, SD, ID, CM, TW, CB, JC, JP, FH, SH

Visualisation: AG, HC

Writing - original draft: AG, HC, BMK

Writing - review & editing: ALL

Information governance: CB BG AM

## Guarantor

BG is the guarantor.

## References

1. Agarwal, A. et al. A living WHO guideline on drugs for covid-19. BMJ 370, m3379 (2020).

2. Hospitalized adults: Therapeutic management. COVID-19 Treatment Guidelines https://www.covid19treatmentguidelines.nih.gov/management/clinical-management/hospitalized-adults--therapeutic-management/.

3. Recommendations | COVID-19 rapid guideline: managing COVID-19 | Guidance | NICE.

4. Department of Health and Social Care. Government launches COVID-19 Antivirals Taskforce to roll out innovative home treatments this autumn. GOV.UK https://www.gov.uk/government/news/government-launches-covid-19-antivirals-taskforce-to-roll-out-innovative-home-treatments-this-autumn(2021).

5. Department of Health and Social Care. UK’s most vulnerable people to receive life-saving COVID-19 treatments in the community. GOV.UK https://www.gov.uk/government/news/uks-most-vulnerable-people-to-receive-life-saving-covid-19-treatments-in-the-community (2021).

6. Population health: COVID-19 treatment methodology. NHS Digital https://digital.nhs.uk/coronavirus/treatments/methodology.

7. England, N. H. S. Interim clinical commissioning policy: neutralising monoclonal antibodies or antivirals for non-hospitalised patients with COVID-19. NHS England https://www.england.nhs.uk/coronavirus/documents/c1603-interim-clinical-commissioning-policy-antivirals-or-neutralising-monoclonal-antibodies-for-non-hospitalised-patients-with-covid-19-version-5/ (2022).

8. Curtis, H. J. et al. Trends and clinical characteristics of 57.9 million COVID-19 vaccine recipients: a federated analysis of patients’ primary care records in situ using OpenSAFELY. Br. J. Gen. Pract. (2021) doi:10.3399/BJGP.2021.0376.

9. Williamson, E. J. et al. Factors associated with COVID-19-related death using OpenSAFELY. Nature 584, 430–436 (2020).

10. Antivirals and nMABs for non-hospitalised COVID-19 patients: coverage report. https://reports.opensafely.org/reports/antivirals-and-nmabs-for-non-hospitalised-covid-19-patients-coverage-report/.

11. National chronic kidney disease audit (NCKDA). LSHTM https://www.lshtm.ac.uk/research/centres-projects-groups/ckdaudit.

12. CAS-ViewAlert. https://www.cas.mhra.gov.uk/ViewandAcknowledgment/ViewAlert.aspx?AlertID=103184.

13. Department for Environment & Affairs, R. 2011 Rural Urban Classification. (2013).

14. Englnad, N. H. S. Safeguarding Adults. https://www.england.nhs.uk/wp-content/uploads/2017/02/adult-pocket-guide.pdf.

15. COVID Medicine Delivery Unit directory. NHS Digital https://digital.nhs.uk/coronavirus/covid-medicine-delivery-unit-directory?key=h58vqkRUup40o27K04xOrtfh7ZXqwRQoOLhXTkGWlbOrVSkwzfTeetw39uGFlc28.

16. antibody-and-antiviral-deployment github repo. (Github).

17. Bhaskaran, K. et al. HIV infection and COVID-19 death: a population-based cohort analysis of UK primary care data and linked national death registrations within the OpenSAFELY platform. Lancet HIV 8, e24–e32 (2021).

18. UK Summary of COVID-19 in the UK. UK Government https://coronavirus.data.gov.uk/.

19. BETA – Data Security Standards - NHS Digital. NHS Digital https://digital.nhs.uk/about-nhs-digital/our-work/nhs-digital-data-and-technology-standards/framework/beta---data-security-standards.

20. Data Security and Protection Toolkit - NHS Digital. NHS Digital https://digital.nhs.uk/data-and-information/looking-after-information/data-security-and-information-governance/data-security-and-protection-toolkit.

21. ISB1523: Anonymisation Standard for Publishing Health and Social Care Data - NHS Digital. NHS Digital https://digital.nhs.uk/data-and-information/information-standards/information-standards-and-data-collections-including-extractions/publications-and-notifications/standards-and-collections/isb1523-anonymisation-standard-for-publishing-health-and-social-care-data.

22. Secretary of State for Health and Social Care - UK Government. Coronavirus (COVID-19): notification to organisations to share information. https://web.archive.org/web/20200421171727/ https://www.gov.uk/government/publications/coronavirus-covid-19-notification-of-data-controllers-to-share-information (2020).

