## Supplementary material for "Trends, variation and clinical characteristics of recipients of antivirals and neutralising monoclonal antibodies for non-hospitalised COVID-19: a descriptive cohort study of 23.4 million people in OpenSAFELY"

***Table S1 Details on how the patient cohorts considered at highest risk from COVID-19 and to be prioritised for treatment with antivirals and nMABs were defined by NHS Digital, and how they were applied in the present study.*** *We include links to the codelists used and describe limitations.*

| **COVID-19 therapeutics high risk cohort identification**<br>(taken from https://digital.nhs.uk/coronavirus/treatments/methodology/rule-logic#chapter-index) | | | | **Codelists** | | **Limitations** |
|---|---|---|---|---|---|---|
| **Cohort** | **Description** | **Data used** | **Rule logic** | **Source** | **OpenCodelists link** | |
| **Down's syndrome** | All patients with down's syndrome | GPES At Risk v4<br><br>HES (2006+) | As per Shielded Patient List rules | COVID-19 therapeutics code list and rule set dataset<br>(Down's syndrome tab from COVID+methodology+cohorts++.xlsx) | https://www.opencodelists.org/codelist/nhsd/downs-syndrome-snomed-ct/3bb246d7/<br><br>https://www.opencodelists.org/codelist/nhsd/downs-syndrome-icd-10/22a2d3a2/ | |
| **Patients with a solid cancer** | Active metastatic cancer and active solid cancers (at any stage) | Non-digital solution via the oncology units | | | https://www.opencodelists.org/codelist/opensafely/lung-cancer-snomed/2020-04-15/<br><br>https://www.opencodelists.org/codelist/opensafely/cancer-excluding-lung-and-haematological-snomed/2020-04-15/ | As non digital solutions were used alongside datasets not available in OpenSAFELY, patients with a solid cancer were identified using a combination of three OpenSAFELY codelists:<br><br>1. Lung Cancer<br>2. Other Cancers<br>3. Chemotherapy or Radiotherapy |
| | All patients receiving chemotherapy within the last 3 months | SACT classified as category B or C in the Chemotherapy Treatment Benchmark Groups as defined by the updated QCOVID Population Risk Assessment code set within the last 12 months | SACT is 3-6 months in arrears, a non-digital solution will cover the deficit via the oncology units. | | https://www.opencodelists.org/codelist/opensafely/chemotherapy-or-radiotherapy-snomed/2020-04-15/ | |
| | Patients receiving radiotherapy within the last 6 months | RTDS within the last 6 months<br><br>HES (2006+)<br>within the last 6 months | RTDS is 3-6 months in arrears, HES data identifies 50% 0-6 months. A non-digital solution will cover the deficit via the oncology units. | | | |
| **Patients with a haematological disease and stem cell transplant recipients** | Allogeneic haematopoietic stem cell transplant (HSCT) recipients in the last 12 months or active graft vs host disease (GVHD) regardless of time from transplant<br><br>Autologous HSCT recipients in the last 12 months (including HSCT for non-malignant diseases) | GPES At Risk v4<br><br>HES (2006+)<br><br>OPCS4 | As per Shielded Patient List rules<br><br>All HSCT transplants are currently included for clinical triage regardless of time from transplant. | 3rd primary dose cohort identification specification<br>(ID 5 - Haematological Transplants set) | https://www.opencodelists.org/codelist/nhsd/haematopoietic-stem-cell-transplant-snomed/3ec3ac16/<br><br>https://www.opencodelists.org/codelist/nhsd/haematopoietic-stem-cell-transplant-icd-10/395e2c2a/<br><br>https://www.opencodelists.org/codelist/nhsd/haematopoietic-stem-cell-transplant-opcs4/204eb8f5/ | Includes non-digital identification so not all patients will be able to be identified.<br><br>Some of the HES codelist have been removed as they don't exist in ICD-10. These are reflected in the codelists available on opencodelists.org. |

| **COVID-19 therapeutics high risk cohort identification** (taken from https://digital.nhs.uk/coronavirus/treatments/methodology/rule-logic#chapter-index) | | | | **Codelists** | | **Limitations** |
|---|---|---|---|---|---|---|
| **Cohort** | **Description** | **Data used** | **Rule logic** | **Source** | **OpenCodelists link** | |
| **Patients with a haematological disease and stem cell transplant Recipients** (continued) | Individuals with haematological malignancies who have<br>• received chimaeric antigen receptor (CAR)-T cell therapy in the last 24 months, or<br>• anti-CD20 monoclonal antibody therapy in the last 12 months<br>• anti-CD38 monoclonal antibody or B-cell maturation agent (BCMA) targeted therapy in the last 6 months | GPES At Risk v4<br><br>HES (2006+)<br><br>OPCS4 | Population Risk Assessment (PRA) code clusters. The current cohorting rules limit Haematological malignancies to:<br>• Chronic lymphocytic leukaemia (CLL)<br>• B-cell lymphoma<br>• Follicular lymphoma<br>• Waldenstrom's macroglobulinaemia<br>• Multiple myeloma<br>• Acute lymphoblastic leukaemia<br>• A non-digital solution will cover the deficit via the oncology units<br><br>There is also a digital cohorting limitation that hospital-only medication is not stored centrally, therefore all the Haematological Malignancies listed above are included for clinical triage, regardless of treatment. | 3rd primary dose cohort identification specification (ID 1 - Haematological Cancers set) | https://www.opencodelists.org/codelist/nhsd/haematological-malignancies-snomed/31a49191/<br><br>https://www.opencodelists.org/codelist/nhsd/haematological-malignancies-icd-10/073f45c0/ | |
| | Individuals with<br>• chronic B-cell lymphoproliferative disorders, receiving systemic treatment or radiotherapy within the last 3 months<br>• reduced peripheral B cell counts or hypogammaglobulinem<br>• Other B-cell lymphoproliferative disorders | GPES At Risk v4<br><br>HES (2006+)<br><br>OPCS4 | Population Risk Assessment (PRA) code clusters<br><br>All chronic B-cell lymphoproliferative disorders are currently included for clinical triage, regardless of time treatment | | | |
| | All patients with sickle cell disease | GPES At Risk v4<br><br>HES (2006+) | As per Shielded Patient List rules | COVID-19 therapeutics code list and rule set dataset ('Sickle' tabs from COVID+methodology+cohorts++.xlsx) | https://www.opencodelists.org/codelist/nhsd/sickle-spl-atrisk v4-snomed-ct/7083ed37/<br><br>https://www.opencodelists.org/codelist/nhsd/sickle-spl-hes-icd-10/789c2d19/ | |

Trends, variation and clinical characteristics of recipients of antivirals and neutralising monoclonal antibodies for non-hospitalised COVID-19: a descriptive cohort study of 23.4 million people in OpenSAFELY

| **COVID-19 therapeutics high risk cohort identification**<br>(taken from https://digital.nhs.uk/coronavirus/treatments/methodology/rule-logic#chapter-index) | | | | **Codelists** | | **Limitations** |
|---|---|---|---|---|---|---|
| **Cohort** | **Description** | **Data used** | **Rule logic** | **Source** | **OpenCodelists link** | |
| **Patients with renal disease** | Renal transplant recipients (including those with failed transplants within the past 12 months), particularly those who<br>● Received B cell depleting therapy within the past 12 months (including alemtuzumab, rituximab<br>● Have an additional substantial risk factors<br>● Have not been vaccinated prior to transplantation | GPES At Risk v4 HES (2006+) OPCS4 | As per Shielded Patient List rules<br><br>Includes all solid transplant patients for clinical triage, regardless of treatment, risk factors and vaccination status, excluding the SPL rule that restricted identification of patients to those with PCPM immunosuppression medication.<br><br>Excludes autologous transplants | | | As the suggested transplant codelists to use here include ALL types of transplants, renal transplant patients will be excluded from this group and included in the solid organ transplant group below.<br><br>Includes non-digital identification of CKD stage 4.<br><br>The ICD-10 code for CKD stage 4 seems to meet the inclusion criteria however it is not included in the definition.<br><br>CKD ICD-10 codes for stages 1-4 are 'excluded' in the criteria. No logic has been included for specific exclusion of ppl with these codes, however they have not been included in the CKD 5 codelist (the inclusion criteria for the eligible cohort).<br><br>As a result of all of the above, this group is CKD stage 5 only. |
| | Non-transplant patients who have received a comparable level of immunosuppression | GPES At Risk v4<br><br>GDPPR<br><br>HES (2006+) | Population Risk Assessment (PRA) code clusters for Immunosuppressant drugs. Anyone, regardless of clinical condition, who has had one of these drugs issued within the last 6 months.<br><br>There is a digital cohorting limitation that hospital-only medication is not stored centrally, therefore there is a non-digital solution for any patients who have had immunosuppressant drugs within the last 6 months that are only hospital prescribed (see section below). | | | |
| | Patients with chronic kidney stage (CKD) 4 (an eGFR less than 30 ml/min/1.73m2) without immunosuppression | Non-digital solution | | | | |
| | Patients with chronic kidney stage (CKD) 5 (an eGFR less than 15 ml/min/1.73m2) without immunosuppression | GPES At Risk v4<br><br>HES (2006+)<br><br>OPCS4 | As per Shielded Patient List rules | COVID-19 therapeutics code list and rule set dataset<br>(CKD Stage 5 tab from COVID+methodology+cohorts++.xlsx) | https://www.opencodelists.org/codelist/nhsd/ckd-stage-5-snomed-ct/2808538e/<br>https://www.opencodelists.org/codelist/nhsd/ckd-stage-5-icd-10/7bad9258/ | |

| **COVID-19 therapeutics high risk cohort identification** (taken from https://digital.nhs.uk/coronavirus/treatments/methodology/rule-logic#chapter-index) | | | | **Codelists** | | **Limitations** |
|---|---|---|---|---|---|---|
| **Cohort** | **Description** | **Data used** | **Rule logic** | **Source** | **OpenCodelists link** | |
| **Patients with liver disease** | Patients with cirrhosis Child's-Pugh class B and C (decompensated liver disease).<br><br>Patients with cirrhosis Child's-Pugh class A who are not on immune suppressive therapy (compensated liver disease) | GPESv4<br><br>HES (2006+) | Clinically Extremely Vulnerable (CEV) codes that were identified for SPL implementation | COVID-19 therapeutics code list and rule set dataset (Liver Cirrhosis tab from COVID+methodology+cohorts++.xlsx) | https://www.opencodelists.org/codelist/nhsd/liver-cirrhosis-icd-10/3e6506cd/<br><br>https://www.opencodelists.org/codelist/nhsd/liver-cirrhosis/64f239d0/ | As the suggested codelists for "Liver patients on immunosuppressive therapy (including patients with and without liver cirrhosis)" is the same as the one used to identify IMID patients below. Liver disease patients meeting this criteria may therefore end up being included in the IMID cohort and not in the 'Liver disease' cohort.<br><br>Includes non-digital identification. |
| | Patients with a liver transplant | GPRSv4<br><br>HES (2006+)<br><br>OPCS4 | As per Shielded Patient List rules<br><br>Includes all solid transplant patients for clinical triage, excluding the SPL rule that restricted identification of patients to those with PCPM immunosuppression medication.<br><br>Excludes autologous transplants | | | |
| | Liver patients on immunosuppressive therapy (including patients with and without liver cirrhosis) | GPES At Risk v4<br><br>GDPPR<br><br>HES (2006+) | Population Risk Assessment (PRA) code clusters Immunosuppressant drugs. Anyone, regardless of clinical condition, who has had one of these drugs issued within the last 6 months.<br><br>There is a digital cohorting limitation that hospital only medication is not stored centrally, therefore there is a non-digital solution for any patients who have had immunosuppressant drugs within the last 6 months that are only hospital prescribed (see section below) | | | |

Trends, variation and clinical characteristics of recipients of antivirals and neutralising monoclonal antibodies for non-hospitalised COVID-19: a descriptive cohort study of 23.4 million people in OpenSAFELY

<table>
<tr><th colspan="4">COVID-19 therapeutics high risk cohort identification<br>(taken from https://digital.nhs.uk/coronavirus/treatments/methodology/rule-logic#chapter-index)</th><th colspan="2">Codelists</th><th rowspan="2">Limitations</th></tr>
<tr><th>Cohort</th><th>Description</th><th>Data used</th><th>Rule logic</th><th>Source</th><th>OpenCodelists link</th></tr>
<tr><td rowspan="2">Patients with immune-mediated inflammatory disorders (IMID)</td><td>IMID with<br>• treated with rituximab or other B cell depleting therapy in the last 12 months<br>• active or unstable disease on corticosteroids, cyclophosphamide, tacrolimus, cyclosporin or mycophenolate.<br>• stable disease on either tacrolimus, cyclophosphamide, cyclosporin or mycophenolate.<br>• active/unstable disease including those on biological monotherapy and on combination biologicals with thiopurine or methotrexate<br>• stable disease on either tacrolimus, cyclophosphamide, cyclosporin or mycophenolate.</td><td>GPES At Risk v4<br><br>GDPPR<br><br>HES (2006+)</td><td>Population Risk Assessment (PRA) code clusters for Immunosuppressant drugs. Anyone, regardless of clinical condition, who has had one of these drugs issued within the last 6 months.<br><br>There is a digital cohorting limitation that hospital only medication is not stored centrally, therefore there is a non-digital solution for any patients who have had immunosuppressant drugs within the last 6 months that are only hospital prescribed (see section below)</td><td>COVID-19 therapeutics code list and rule set dataset (Immunosuppressant Drugs (PRA) tab from COVID+methodology+cohorts++.xlsx)</td><td>https://www.opencodelists.org/codelist/nhsd/immunosuppressant-drugs-pra-dmd/526d9f5f/<br><br>https://www.opencodelists.org/codelist/nhsd/immunosuppressant-drugs-pra-snomed/6b7d1294/</td><td>Includes non-digital identification<br><br>Within the NHSD DM+D codelist for Immunosuppressant drugs, 3 SNOMED codes were identified. These codes are deprecated SNOMED codes and have been put into a separate SNOMED codelist which was used in the study in combination with the DM+D codelist..</td></tr>
<tr><td>• IMID with active/unstable disease on corticosteroids<br>• IMID with stable disease on corticosteroids<br>• IMID with stable disease on corticosteroids</td><td>GPES At Risk v4<br><br>GDPPR</td><td>All patients on corticosteroids (where 2 prescriptions have been issued in 3 month, or 4 prescriptions in 12 months) are included in the for clinical triage.</td><td>COVID-19 therapeutics code list and rule set dataset Oral Steroid Drugs (PRA) tab from COVID+methodology+cohorts++.xlsx)</td><td>https://www.opencodelists.org/codelist/nhsd/oral-steroid-drugs-pra-dmd/44292c02/<br><br>https://www.opencodelists.org/codelist/nhsd/oral-steroid-drugs-snomed/1ae93909/</td><td>Within the NHSD DM+D codelist for Oral Steroid drugs, 1 SNOMED code was identified. This code is a deprecated SNOMED code. The code has been put into a separate the SNOMED codelist which was used in the study in combination with the DM+D codelist..</td></tr>
</table>

Trends, variation and clinical characteristics of recipients of antivirals and neutralising monoclonal antibodies for non-hospitalised COVID-19: a descriptive cohort study of 23.4 million people in OpenSAFELY

| **COVID-19 therapeutics high risk cohort identification**<br>(taken from https://digital.nhs.uk/coronavirus/treatments/methodology/rule-logic#chapter-index) | | | | **Codelists** | | **Limitations** |
|---|---|---|---|---|---|---|
| **Cohort** | **Description** | **Data used** | **Rule logic** | **Source** | **OpenCodelists link** | |
| Primary immune deficiencies | • Common Variable Immunodeficiency (CVID)<br>• Undefined primary antibody deficiency on immunoglobulin (or eligible for Ig)<br>• Hyper-IgM syndromes<br>• Good's syndrome (thymoma plus B-cell deficiency)<br>• Severe Combined Immunodeficiency (SCID)<br>• Autoimmune polyglandular syndromes/autoimmune polyendocrinopathy, candidiasis, ectodermal dystrophy (APECED syndrome)<br>• Primary immunodeficiency associated with impaired type I interferon signalling<br>• X-linked agammaglobulinaemia (and other primary agammaglobulinaemias) | GPES At Risk v4<br><br>GDPPR<br><br>HES (2006+) | Population Risk Assessment (PRA) code clusters<br><br>There is a digital cohorting limitation in that Autoimmune polyglandular syndromes/autoimmune polyendocrinopathy, candidiasis, ectodermal dystrophy (APECED syndrome) is not included. A non-digital solution will cover the deficit via the specialist units (see section below) | COVID-19 therapeutics code list and rule set dataset (Immunosupression (PCDCluster) tab from COVID+methodology+cohorts++.xlsx) | https://www.opencodelists.org/codelist/nhsd/immunosupression-pcdcluster-snomed-ct/09936066d/ | Includes non-digital identification |
| HIV/AIDS | Patients with high levels of immune suppression, have uncontrolled/untreated HIV (high viral load) or present acutely with an AIDS defining diagnosis<br><br>On treatment for HIV with CD4 <350 cells/mm3 and stable on HIV treatment or CD4>350 cells/mm3 and additional risk factors (e.g. age, diabetes, obesity, cardiovascular, liver or renal disease, homeless, those with alcohol-dependence) | GPES At Risk v4<br><br>HES (2006+)<br><br>OPCS4 | Population Risk Assessment (PRA) code clusters Includes all people with HIV regardless of CD4 count, viral load and risk factors. | 3rd primary dose cohort identification specification (ID 3 - Immunosuppression code set) | https://www.opencodelists.org/codelist/nhsd/hiv-aids-snomed/68ca529e/<br><br>https://www.opencodelists.org/codelist/nhsd/hiv-aids-icd10/01d9c5d4/ | |

| | | | | | | |
|---|---|---|---|---|---|---|
| Solid organ transplant recipients | All recipients of solid organ transplants not otherwise specified above | GPES At Risk v4<br><br>HES (2006+)<br><br>OPCS4 | Clinically Extremely Vulnerable codes that were identified for SPL implementation | COVID-19 therapeutics code list and rule set dataset (Transplant tabs from COVID+methodology+cohorts++.xlsx) | https://www.opencodelists.org/codelist/nhsd/transplant-spl-atriskv4-snomed-ct/57747a02/<br><br>https://www.opencodelists.org/codelist/nhsd/transplant-spl-hes-opcs4/1b47de52/<br><br>https://www.opencodelists.org/codelist/nhsd/transplant-conjunctiva-spl-hes-opcs4/2dcc78c3/<br><br>https://www.opencodelists.org/codelist/nhsd/transplant-conjunctiva-y-codes-spl-hes-opcs4/14bd058e/<br><br>https://www.opencodelists.org/codelist/nhsd/transplant-ileum_1-y-codes-spl-hes-opcs4/01131245/<br><br>https://www.opencodelists.org/codelist/nhsd/transplant-ileum_1-spl-hes-opcs4/3331f8af/<br><br>https://www.opencodelists.org/codelist/nhsd/transplant-ileum_2-y-codes-spl-hes-opcs4/68039f0f<br><br>https://www.opencodelists.org/codelist/nhsd/transplant-ileum_2-spl-hes-opcs4/1a22857a/<br><br>https://www.opencodelists.org/codelist/nhsd/transplant-replacement-of-organ-spl-hes-opcs4/46dbebf8/<br><br>https://www.opencodelists.org/codelist/nhsd/transplant-stomach-spl-hes-opcs4/6550df19/ | The NHS Digital solid organ transplant list covers all transplants (including renal and liver) and ALL transplant patients will be included in this group and not any of the other condition subgroups, as described by NHS Digital.<br><br>Some HES transplant codes contain 2 codes required to identify transplants. These have been included with logic in the study definition, however for each '2 code' transplant, the component codelists were added to opencodelists. For example; stomach transplant is defined as having G388: 'Other specified open operations on the stomach' + any of the 'replacement of organ' Y codes (for example Y018 'Other specified replacement of organ'). The date of the Y-code, defining the nature of the procedure as a transplant is used as a date restriction in looking for the organ specific code, thus both codes need to be present on the same hospital spell admission date to qualify as a transplant.<br><br>The code Z94 is described as a transplant specific OPCS code in the NHSD source, however this OPCS code actually describes the laterality of a procedure. This code was removed from the analyses as it led to incorrect over identification, and is assumed to be an error in the published codelist. |

Trends, variation and clinical characteristics of recipients of antivirals and neutralising monoclonal antibodies for non-hospitalised COVID-19: a descriptive cohort study of 23.4 million people in OpenSAFELY

| **COVID-19 therapeutics high risk cohort identification** (taken from https://digital.nhs.uk/coronavirus/treatments/methodology/rule-logic#chapter-index) | | | | **Codelists** | | **Limitations** |
|---|---|---|---|---|---|---|
| **Cohort** | **Description** | **Data used** | **Rule logic** | **Source** | **OpenCodelists link** | |
| Rare neurological conditions | Multiple sclerosis | GPES At Risk v4<br><br>GDPPR<br><br>HES (2006+) | Population Risk Assessment (PRA) code clusters collated by the nMABS clinicians to restrict the data to the 4 disease groups. | COVID-19 therapeutics code list and rule set dataset (Multiple Sclerosis, Motor neurone disease, Myasthenia Gravis and Huntingtons diesease tabs from COVID+methodology+cohorts++.xlsx) | https://www.opencodelists.org/codelist/nhsd/multiple-sclerosis/4be2c69b/<br><br>https://www.opencodelists.org/codelist/nhsd/multiple-sclerosis-snomed-ct/7336ad2d/ | |
| | Motor neurone disease | | | | https://www.opencodelists.org/codelist/nhsd/motor-neurone-disease-icd-10/4117c6c3/<br><br>https://www.opencodelists.org/codelist/nhsd/motor-neurone-disease-snomed-ct/5a2739f8/ | |
| | Myasthenia gravis | | | | https://www.opencodelists.org/codelist/nhsd/myasthenia-gravis/32d35366/<br><br>https://www.opencodelists.org/codelist/nhsd/myasthenia-gravis-snomed-ct/0c462063/ | |
| | Huntington’s disease | | | | https://www.opencodelists.org/codelist/nhsd/huntingtons/7e01ad05/<br><br>https://www.opencodelists.org/codelist/nhsd/huntingtons-snomed-ct/25559398/ | |

***Table S2  COVID-19 treatment specific eligibility and exclusion criteria, as per the Interim Clinical Commissioning Policy (published on 28 January 2022, effective from 10 February)*[1] *for non-hospitalised COVID-19 patients, and how these were applied in the present study.*** *These criteria were applied to identify potential breaches in guidance for patients who received each type of treatment.*

| Treatment | Eligibility/Exclusion Criteria | | Criteria applied in present study* |
|---|---|---|---|
| **Paxlovid** | **Eligibility criteria** | Clinical judgement deems that an antiviral is the preferred option | Not possible[†1] |
| | | Treatment is commenced within 5 days of symptom onset (extended up to a maximum of 7 days if clinically indicated) | As it was not possible to accurately define symptom onset date, an individual's positive SARS-CoV-2 test date was used as a proxy and used a maximum limit of 5 days. |
| | | The patient does NOT have a history of advanced decompensated liver cirrhosis or stage 3-5 chronic kidney disease | Not a member of the renal disease of liver disease group as defined in Table S1. |
| | | Paxlovid has been deemed safe following guidance from the appropriate specialty team(s) | Not possible[†1] |
| | **Exclusion criteria** | Children aged less than 18 years | Age>=18 as of earliest of treatment date or positive SARS-CoV-2 test date. |
| | | Pregnancy | Not possible[†2] |
| | | The patient is taking any of the contraindicated medications | Not possible[†1] |
| **Sotrovimab** | **Eligibility criteria** | Clinical judgement deems that an nMAB is the preferred option | Not possible[†1] |
| | | Treatment is delivered within 5 days of symptom onset (extended up to a maximum of 7 days if clinically indicated) | As it was not possible to accurately define symptom onset date, an individual's positive SARS-CoV-2 test date was used as a proxy and used a maximum limit of 5 days. |
| | **Exclusion criteria** | Children aged under 12 years | Age>=12 as of earliest of treatment date or positive SARS-CoV-2 test date. |

| Treatment | Eligibility/Exclusion Criteria | | Criteria applied in present study* |
|---|---|---|---|
| | | Adolescents (aged 12-17) weighing 40kg and under | Age 12-17 as of earliest of treatment date or positive SARS-CoV-2 test date, andL latest available weight>40kg, as recorded on or before treatment date or positive SARS-CoV-2 test date using this SNOMED codelist. |
| **Remdesivir** | **Eligibility criteria** | Clinical judgement deems that an antiviral is the preferred option | Not possible[†] |
| | | Treatment with PF-07321332 (nirmatrelvir) plus ritonavir is contraindicated or not possible | Not possible[†] |
| | | Treatment is commenced within 7 days of symptom onset | As it was not possible to accurately define symptom onset date, positive SARS-CoV-2 test date was used as a proxy and used a maximum limit of 7 days. |
| | **Exclusion criteria** | Children aged under 12 years | Age>=12 as of positive SARS-CoV-2 test date. |
| | | Adolescents (aged 12-17) weighing 40kg and under | Age 12-17 as of earliest of treatment date or positive SARS-CoV-2 test date and lLatest available weight>40kg, as recorded on or before each patient's treatment date or positive SARS-CoV-2 test date using this SNOMED codelist. |
| **Molnupiravir** | **Eligibility criteria** | Treatment Paxlovid AND sotrovimab are contraindicated or not possible | Not possible[†1] |
| | | Treatment is commenced within 5 days of symptom onset (extended up to a maximum of 7 days if clinically indicated) | As it was not possible to accurately define symptom onset date, positive SARS-CoV-2 test date was used as a proxy and used a maximum limit of 5 days. |
| | **Exclusion criteria** | Children aged less than 18 years | Age>=18 as of earliest of treatment date or positive SARS-CoV-2 test date. |
| | | Pregnancy | Not possible[†2] |

** Note: all patients who received treatment were included in the population even if they were not identified as meeting the initial eligibility criteria, as described in Table 1*

[†1] *It was not possible to identify* any forms of clinical judgement that would have informed treatment options

[†2] *Not currently applied due to limitations and inaccuracies in using pregnancy codes as they are not always well-captured in GP records. Methods are being validated and will be applied in future analyses if appropriate*

Trends, variation and clinical characteristics of recipients of antivirals and neutralising monoclonal antibodies for non-hospitalised COVID-19: a descriptive cohort study of 23.4 million people in OpenSAFELY

***Figure S1 Weekly proportion of eligible patients receiving an antiviral or nMAB for treating COVID-19 since 11th December 2021, stratified by high risk cohort.*** *The x-axis represents the eligibility week of patients with weekly proportions calculated as the number of weekly eligible patients who went on to receive treatment divided by the number of weekly eligible patients.*

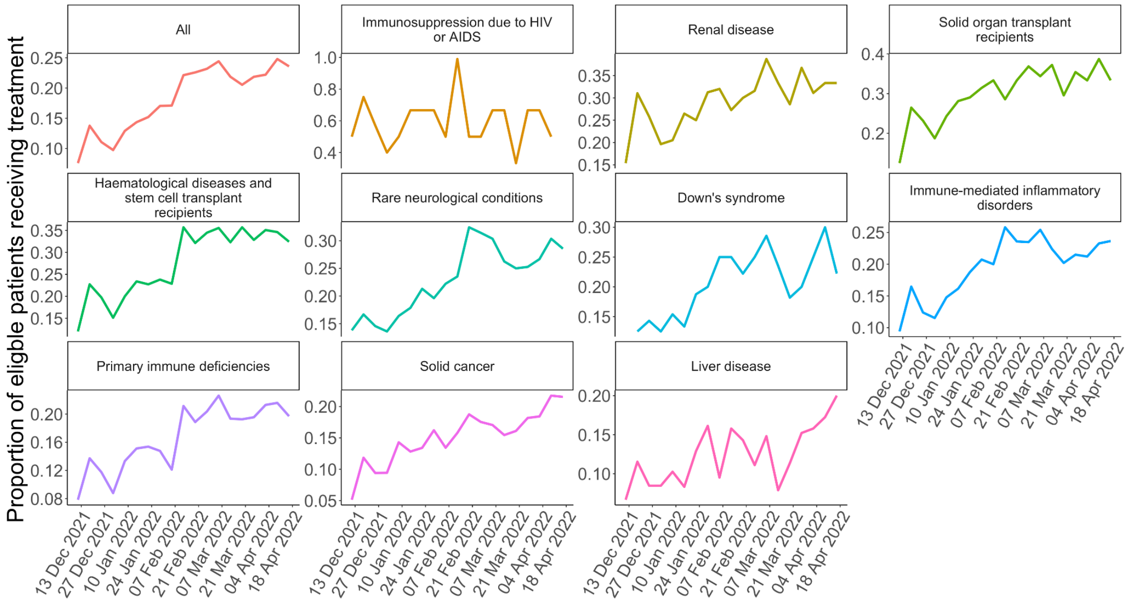


***Figure S2 Breakdown of possible inconsistencies with guidance on eligibility and exclusion criteria in treated COVID-19 patients.*** *Treatment eligibility window for Paxlovid, sotrovimab and molnupiravir was 5 days from positive SARS-CoV-2 test (used as a proxy for symptom onset date) and 7 days for remdesivir.*

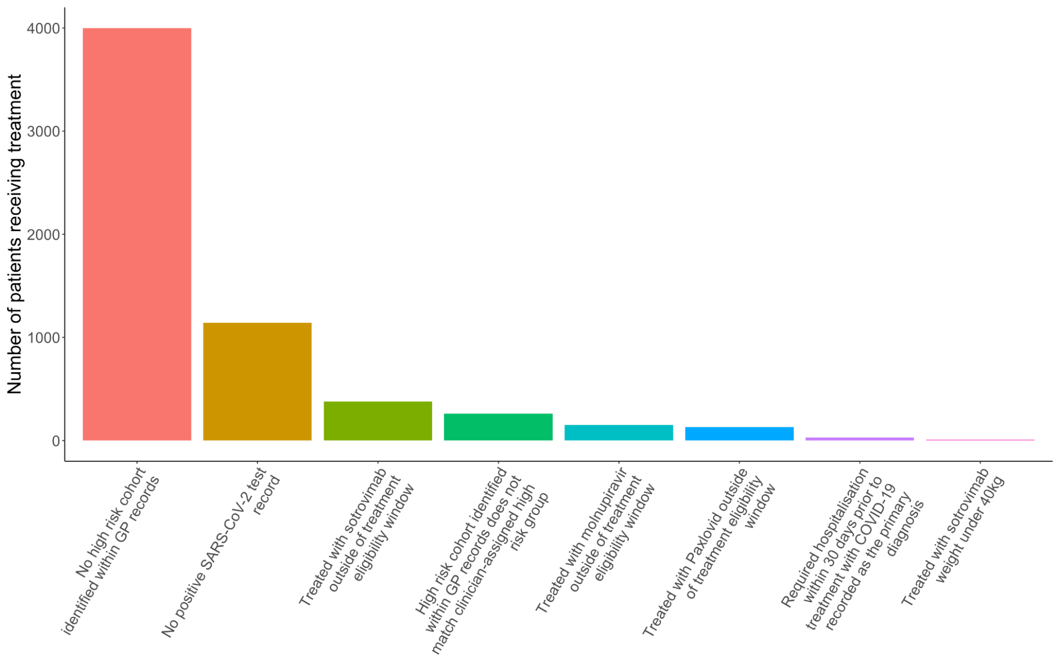

Trends, variation and clinical characteristics of recipients of antivirals and neutralising monoclonal antibodies for non-hospitalised COVID-19: a descriptive cohort study of 23.4 million people in OpenSAFELY

***Figure S3 Time (number of days) between positive SARS-CoV-2 test and treatment for COVID-19, broken down by treatment type.*** *Treatment types are restricted to the three most commonly used treatments.*

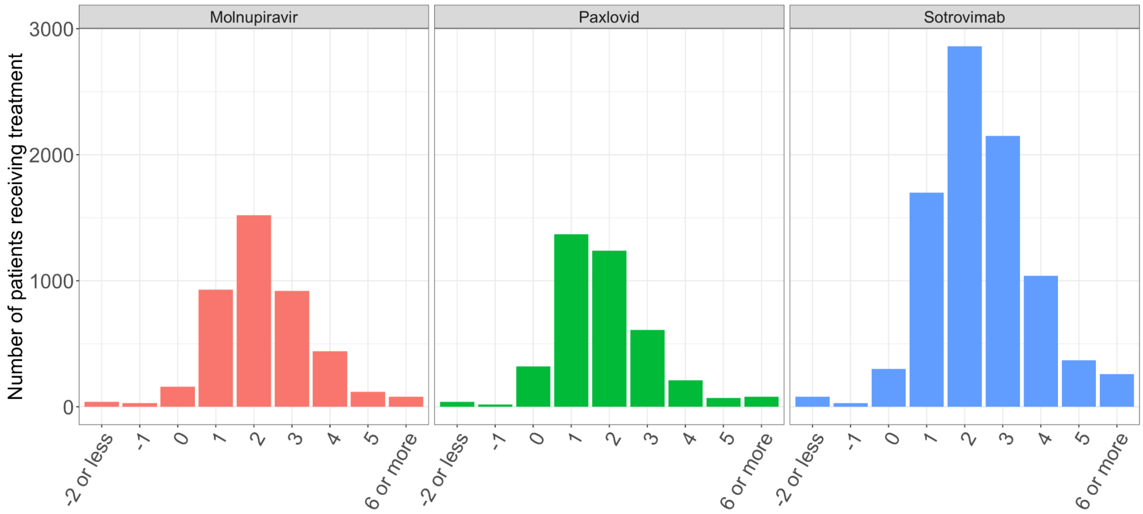


**Figure S4 Time between positive SARS-CoV-2 test and treatment for COVID-19, broken down by high risk cohort**

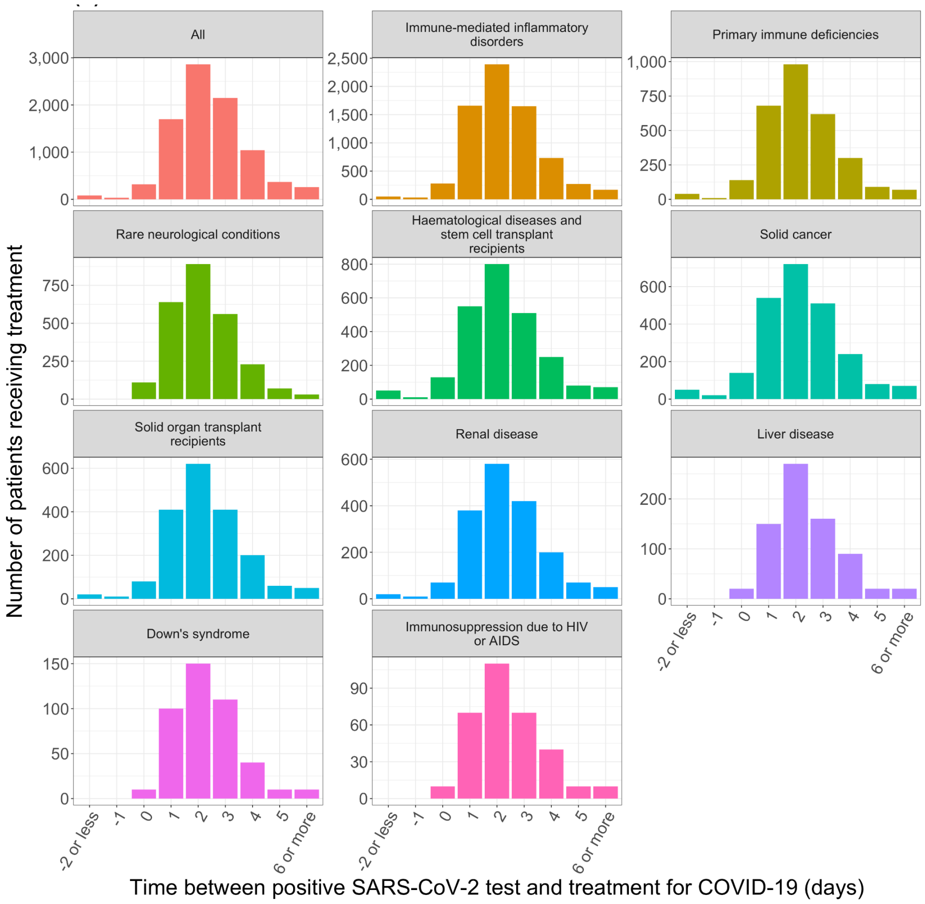

**Figure 3 Weekly proportion of eligible patients receiving an antiviral or nMAB for treating COVID-19 since 11th December 2021, stratified by STP decile.**

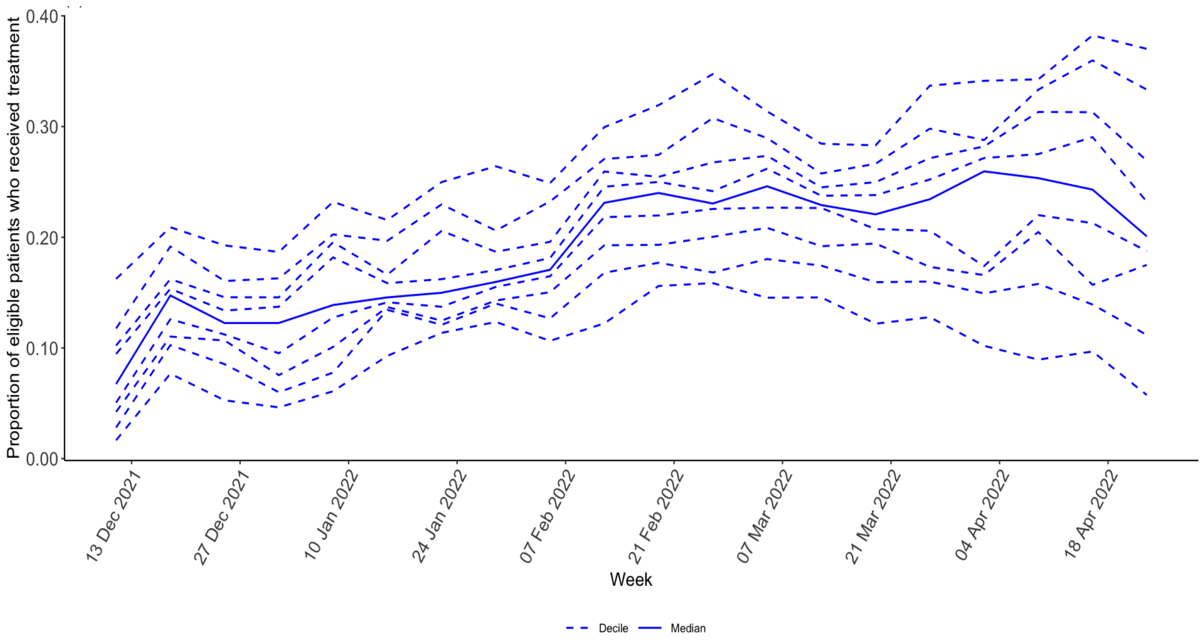


### REFERENCES

1. NHS England. Interim clinical commissioning policy: neutralising monoclonal antibodies or antivirals for non-hospitalised patients with COVID-19. *NHS England* https://www.england.nhs.uk/coronavirus/publication/interim-clinical-commissioning-policy-neutralising-monoclonal-antibodies-or-antivirals-for-non-hospitalised-patients-with-covid-19/ (2022).
